# Transitions from smoking to exclusive e-cigarette use, dual use, or stopping nicotine use in ALSPAC and their association with modifiable and sociodemographic factors

**DOI:** 10.1101/2024.10.14.24315457

**Authors:** Alexandria Andrayas, Jon Heron, Jasmine Khouja, Hannah Jones, Hannah Sallis, Marcus R Munafò, Lindsey Hines, Elinor Curnow

## Abstract

**Background:** E-cigarettes can aid smoking cessation and reduce carcinogen exposure. Understanding differences in characteristics between young adults in the UK who quit smoking, with or without e-cigarettes, or who dual use can help identify and tailor interventions.

**Methods:** There were 858 participants in the Avon Longitudinal Study of Parents and Children who reported smoking at 21 years. Transitions to the first reported non-exclusive smoking event were measured at 22-30 years and include stopping nicotine use (not currently smoking nor using e-cigarettes), exclusive e-cigarette use, and dual use (smoking and using e-cigarettes). Associations between these transitions and substance use, sociodemographic, and health characteristics were examined through discrete-time survival analysis. Analyses were adjusted for early life confounders, and weighted to mitigate bias. Multiple imputation was used to analyse partially-observed data.

**Results:** More participants reported stopping nicotine use (52%) than using e-cigarettes (dual use = 27%; exclusive e-cigarette use = 9%). Smoking frequently at age 21 (Subdistribution Hazard Ratio [SHR] = 0.28, CI = 0.22-0.35) and having many friends who smoke at age 21 (SHR = 0.64, CI = 0.50-0.81) decreased the likelihood of stopping nicotine use, and increased the likelihood of dual use (frequent smoking SHR = 3.00, CI = 1.96-4.59; peer smoking SHR = 1.55, CI = 1.07-2.24) and exclusive e-cigarette use (frequent smoking SHR = 2.84, CI = 1.26-6.42). Participants with lower education at age 20 were less likely to stop using nicotine (SHR = 0.68, CI = 0.52-0.90), and more likely to dual use (SHR = 1.72, CI = 1.03-2.90). Cannabis use at age 20 (SHR = 0.67, CI = 0.49-0.92), drug use at age 20 (SHR = 0.77, CI = 0.59-0.99), less exercise at age 18 (SHR = 0.71, CI = 0.53-0.95), and parenthood at age 21 (SHR = 0.46, CI = 0.27-0.79) reduced the likelihood of stopping nicotine use. Higher BMI at age 18 (SHR = 1.58, CI = 1.08-2.31) increased the likelihood of dual use.

**Conclusions:** Young adults who smoke more frequently, have many peers who smoke, have lower education, exercise less, use cannabis or other drugs likely need greater efforts from stop smoking intervention. Those with greater smoking frequency, more friends who smoke or lower education may need additional support to completely switch to e-cigarettes. Others not engaging with e-cigarettes may benefit from targeted interventions to encourage harm reduction.

## Introduction

Tobacco control approaches in the UK have achieved great success given smoking rates have been declining since records began in 1974^1^. However, harm reduction remains essential for people who currently smoke and may have failed, or do not intend, to quit smoking. Electronic cigarettes (e-cigarettes) are a potential harm reduction tool that can help people stop smoking. E-cigarettes are devices that heat liquid, often containing propylene glycol, vegetable glycerin, flavourings, nicotine, and other additives, to form an aerosol which can be inhaled. The aerosol contains fewer and lower levels of harmful chemicals and toxicants compared to cigarette smoke^2^. When used for smoking cessation, nicotine e-cigarettes can be more successful than traditional nicotine replacement therapy (NRT), nicotine-free e-cigarettes, or behavioural support alone^3,4^.

Since emerging on the UK market in 2007^5^, e-cigarettes have grown in popularity. An estimated 4.7 million adults in Great Britain used e-cigarettes in 2023, 56% of whom previously smoked, and 37% of whom currently smoke^6^. Recent reduction in smoking rates may be partly driven by the availability of e-cigarettes, which may attract people who would not have otherwise used NRT^7–10^. E-cigarettes have been regulated as consumer products as part of the UK Tobacco and Related Products Regulations since 2016^11–13^. Recently the UK government and National Health Service began pioneering a Swap to Stop campaign aimed to help people quit smoking by offering free vape starter kits and behavioural support^14^. However, not all people who smoke completely substitute cigarettes for e-cigarettes or use them to try to stop smoking.

When people continue to smoke alongside using e-cigarettes this is referred to as dual use. Dual use is associated with greater nicotine dependence and consumption of both cigarettes and e-cigarettes^15^, may expose a person to similar levels of carcinogens as only smoking^16^, and is unlikely to substantially reduce harm if it does not lead to quitting smoking^17^. Therefore, it is important to understand the factors associated with dual use to target those most in need of support to effectively use e-cigarettes to stop smoking.

Previous research has identified predictors of e-cigarette use, including higher tobacco dependence^18^, smoking by friends or family and higher impulsivity^19^, internalising mental health symptoms^20^, drunkenness, energy drink use, and poor academic achievement^21^, and conduct problems^22^. Studies have also shown that e-cigarette use may be less persistent over time than cigarette use^23–25^, and may facilitate a quicker transition to smoking abstinence^26^. However much of this research has focused on adolescence or the entire adult age range, has limited follow up data up to a few years, and has focused on one or few predictors. This study adds to existing knowledge by utilising 8 years of multiple follow ups in a rich longitudinal dataset of young adults in the UK.

This study investigates differences in a broad range of characteristics between young adults in the UK who transition from smoking to using e-cigarettes exclusively or while smoking (dual use), or quit smoking without using e-cigarettes. Understanding the factors associated with these transitions could help identify target populations who are less likely to quit smoking or use e-cigarettes, or more likely to dual use.

## Methods

### Aims

This study aims to (1) investigate the first self-reported transitions from smoking, to stopping nicotine use, exclusive e-cigarette use, and dual use; and (2) explore the association of different transitions from smoking with baseline substance use, sociodemographic, and health characteristics.

### Participants

Pregnant women resident in Avon, UK with expected delivery dates between 1st April 1991 and 31^st^ December 1992 were invited to take part in the study. The initial number of pregnancies enrolled was 14,541, including 14,203 unique mothers, of which 13,988 children were alive at 1 year of age. The total sample size for analyses using any data collected after the age of seven is 15,447 pregnancies, of which 14,901 children were alive at 1 year of age. A total of 14,833 unique women (G0 mothers) were enrolled in ALSPAC as of September 2021. Detailed information has been collected on these women and their offspring (G1) at regular intervals^27–29^. 12,113 G0 partners have been in contact with the study by providing data or formally enrolling when this started in 2010. 3,807 G0 partners are currently enrolled^30,31^. Study data were collected and managed using REDCap electronic data capture tools hosted at the University of Bristol^32^. The study website contains details of available data through a fully searchable data dictionary and variable search tool: http://www.bristol.ac.uk/alspac/researchers/our-data/.

The eligible sample for the present analyses (n=3,290) consists of G1 participants in ALSPAC who responded to the 21+ questionnaire (administered from mid-December 2013 when participants were approximately 21 years of age) and reported whether or not they had smoked in the past 30 days. The analytic sample (n=858) is limited to participants who responded ‘Yes’ to smoking in the past 30 days. Of these 803 (94%) participants also reported their smoking and e-cigarette use behaviour at least once in a following timepoint. Further details are shown in Supplementary Figure S1.

### Outcomes

Repeated measures of e-cigarette use have been collected in ALSPAC from approximately 22 years of age. At each of five timepoints (22, 23, 24, 28, 30 years [y]), nicotine use was characterised based on self-reported smoking in the past 30 days, and ever or current e-cigarette use, at the time of the questionnaire. At the 30+ questionnaire (at approximately 30 y) participants were asked if they had used e-cigarettes in the past 30 days rather than if they were currently using e-cigarettes, as in previous timepoints.

Transitions from smoking are defined as the first reported, if any, non-exclusive smoking event (*i.e.* whichever occurred first out of stopping nicotine use, exclusive e-cigarette use, or dual use, during the 22 to 30 year time period). The time to each transition was defined as the difference in approximate whole number of years between the 21+ questionnaire and the timepoint at which the event was reported (*i.e.* using discrete-time categories of 1, 2, 3, 7, or 9 years). An illustrative example is shown in Supplementary Figure S2.

#### Stopping nicotine use

Participants who transitioned to “stopping nicotine use” had not smoked in the past 30 days at the time the event was reported, and had either never used e-cigarettes or not reported currently using e-cigarettes at a prior timepoint. It is important to note that we use the term “stopping nicotine use” for conciseness, but this definition refers to stopping the use of nicotine products assessed in this study (*i.e.* cigarettes and e-cigarettes), and does not take into account other sources of nicotine.

#### Exclusive e-cigarette use

Participants who transitioned to “exclusive e-cigarette use” had not smoked in the past 30 days but reported currently using e-cigarettes at the time the event was reported, without reporting stopping nicotine use or using e-cigarettes alongside smoking at a prior timepoint.

#### Dual use

Participants who transitioned to “dual use” had smoked in the past 30 days and reported currently using e-cigarettes at the time the event was reported, without reporting stopping nicotine use or exclusively using e-cigarettes at a prior timepoint.

### Exposures

Eleven dichotomised substance use, sociodemographic, and health characteristics were investigated, spanning a wide range of potential intervention targets. Available measures collected during or prior to the 21+ questionnaire were selected.

#### Substance use

Measures related to substance use were smoking frequency (*occasional vs weekly or more*) at age 21, and binge drinking frequency (*never, monthly, or less vs weekly or more*), past year cannabis use (*never, not in the past year, less than monthly vs monthly or more*), and past year drug use (*no vs yes*), all collected at age 20. Binge drinking refers to drinking six or more units of alcohol.

#### Social and sociodemographic factors

Measures related to social and sociodemographic factors were peer smoking (*none, a few, or some vs most or all*) and educational attainment (*degree-level vs A-level, below, or other*), both collected at age 20, and parenthood (*no vs yes*) and neighbourhood deprivation, estimated via Townsend deprivation scores (*least deprived quintile vs more deprived quintiles*), both collected at age 21. Peer smoking refers to the number of friends who had smoked cigarettes from 18 to 21 years. A-level (“Advanced Level”) qualifications typically follow General Certificate of Secondary Education (GCSE) examinations or equivalent, usually in the age range of 16 to 18 years.

#### Physical and mental health

Measures related to physical and mental health were BMI (*less than 25 vs 25 or more*) and frequency of exercise in the past year (*weekly or more vs less than weekly*), both collected at age 18, and depressive symptoms in the previous 2 weeks, estimated via Short Mood and Feelings Questionnaire (SMFQ) scores *(less than 12 vs 12 or more*) collected at age 21. Higher scores on the SMFQ suggest more severe depressive symptoms, where a score of 12 or higher may indicate the presence of depression.

### Confounders

Analyses were adjusted for four early-life confounders; sex assigned at birth (*male vs female*), ethnicity (*white vs minority ethnic*), average household income (GBP) per week (*560+ vs 430-559, 240-429, <*240) at age 11, and parental smoking (*no vs yes*) reported by mothers and/or partners when the participant was age 12. Although it is best to use precise terminology to describe the specific ethnicity of a person or group, this data was unavailable. Instead ALSPAC derived ethnicity based on the reported ethnic groups of the mother and father. Here we use “minority ethnic” to refer to participants from all other ethnic groups combined when compared to participants with two white parents. This uses ‘minority’ in a UK context but these groups often represent the global majority.

### Statistical analysis

Data were cleaned and variables created using Stata (version 18)^33^ removing participants who had withdrawn their consent by the time data were accessed. Statistical analyses and data visualisation were carried out in R (version 4.3.3)^34^.

Since the first transition from smoking could be any one of three mutually-exclusive events, subdistribution hazard discrete-time models^35^ were used. These can accommodate competing risks in a discrete time setting and were used to investigate associations between the 11 substance use, sociodemographic, and health characteristics being investigated (18-21y) and the first reported transitions from smoking (22-30y). Results describe the relative change in the subdistribution hazard ratio associated with each characteristic adjusted for the four baseline confounders.

The subdistribution hazard function is defined as the instantaneous rate of occurrence, at any time, of the event of interest (where this denotes a particular transition from smoking), in participants who have not yet experienced an event of that type (meaning they either experienced no event, or a competing event *i.e.* any other transition from smoking). Subdistribution hazard models offer inferences about the relative magnitude and direction of the effect of covariates on the cumulative incidence function (CIF) for the event of interest but do not necessarily convey the absolute magnitude of this effect, nor can magnitudes between different subdistribution hazard models be compared. Therefore discussion of these results focuses on the relative importance and direction of effects^36^ using subdistribution hazard ratios (SHR) and 95% confidence intervals (CI).

Multiple imputation by chained equations was used to impute any missing data^37,38^. 100 multiple imputations with 25 iterations were used. Repeated and related measures of the investigated substance use, sociodemographic, and health characteristics were included as auxiliary variables. Inclusion of these into the imputation model was based on a minimum correlation threshold of 0.1. Correlation and predictor matrices are shown in Supplementary Figures S3-6. The first reported transitions from smoking were passively imputed to avoid collinearity issues with the nicotine use categories from which the transitions were derived.

As the analytic sample is conditioned on smoking status and this may induce selection or collider-conditioning bias, a logistic regression model was fitted to predict smoking status at the 21+ questionnaire. Multiple imputation in the eligible sample was used for these weighting purposes. Pooled predicted probabilities were then utilised to derive weights related to selection into the analytic sample of participants who smoke. These weights were accommodated into downstream discrete-time survival analyses^39,40^. This is described further in Supplementary Text S2 and Figure S7.

## Results

### Descriptive statistics

Reported exclusive smoking declined with time, while stopping nicotine use and e-cigarette use increased with time. Initially dual use was more commonly reported than exclusive e-cigarette use, however at later timepoints similar proportions are observed (Supplementary Figure S8). By the end of follow-up 12% of participants were still exclusively smoking and had not reported quitting or using e-cigarettes at any point (Figure 1). Of those with an observed transition from smoking, most moved to stopping nicotine use (52%) and a large proportion of these particular transitions occurred approximately 1 year following the 21+ questionnaire. Movement to using e-cigarettes was seen in 36% of participants and more participants transitioned to dual use (27%) than to exclusive e-cigarette use (9%).

**Figure 1.**
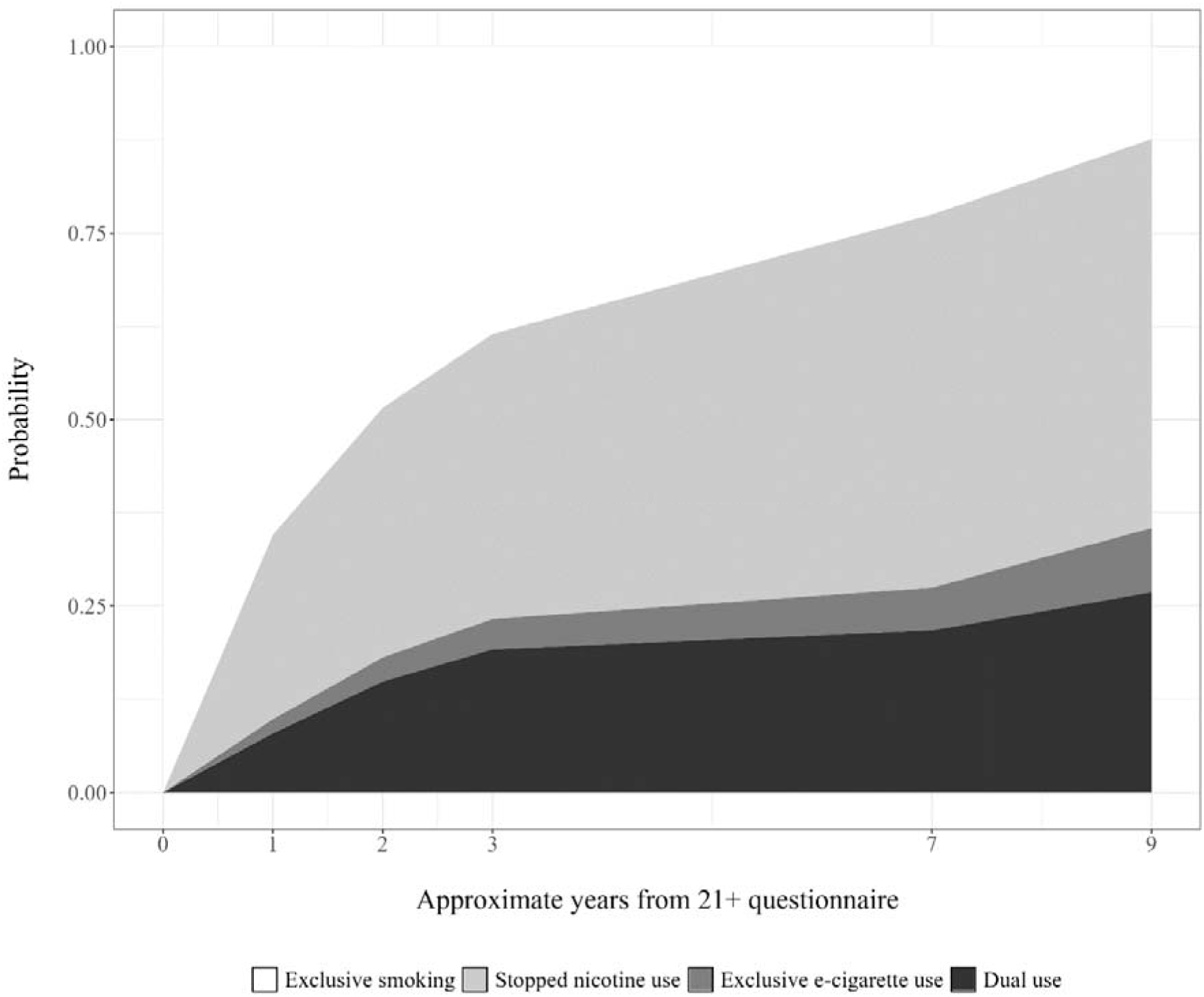
Stacked cumulative incidence curves showing the probability of each first transition from smoking over time since 21+ questionnaire.

In the full analytic sample (n=858) only 8% of participants reported being a parent at 21 years and only 4% had at least one parent from a minority ethnic background (Table 1). Frequency of exercise measured in the 18-year questionnaire showed the greatest amount of missingness (44%) out of the investigated characteristics. Differences in characteristics by each transition from smoking can be found in Supplementary Table S1. Transitions to exclusive e-cigarette use and dual use differed by sex, and transitions to stopping nicotine use and dual use differed by parental smoking at age 12.

**Table 1.** Participant characteristics (n=858).

| Characteristic | Missing<sup>f</sup> | Non-missing<sup>f</sup> |
| --- | --- | --- |
| *Early-life confounders* |  |  |
| **Sex assigned at birth** | 0 (0%) |  |
| Male |  | 308 (36%) |
| Female |  | 550 (64%) |
| **Ethnicity** | 70 (8.2%) |  |
| White |  | 760 (96%) |
| Minority ethnic |  | 28 (3.6%) |
| **Household income (11y<sup>2</sup>)** | 207 (24%) |  |
| 560+ |  | 265 (41%) |
| 430-559 |  | 141 (22%) |
| 240-429 |  | 176 (27%) |
| <240 |  | 69 (11%) |
| **Parental smoking (12y)** | 137 (16%) |  |
| No |  | 547 (76%) |
| Yes |  | 174 (24%) |
| *Substance use* |  |  |
| **Smoking frequency (21y)** | 5 (0.6%) |  |
| Occasional |  | 312 (37%) |
| Weekly or more |  | 541 (63%) |
| **Binge drinking frequency (20y)** | 228 (27%) |  |
| Never, monthly, or less |  | 353 (56%) |
| Weekly or more |  | 277 (44%) |
| **Cannabis use (20y)** | 215 (25%) |  |
| Never, less than monthly, or not in past year |  | 489 (76%) |
| Monthly or more |  | 154 (24%) |
| **Drug use (20y)** | 245 (29%) |  |
| No |  | 347 (57%) |
| Yes |  | 266 (43%) |
| ***Social and sociodemographic factors*** |  |  |
| **Peer smoking (20y)** | 218 (25%) |  |
| None, a few, or some |  | 261 (41%) |
| Most or all |  | 379 (59%) |
| **Education (20y)** | 211 (25%) |  |
| Degree-level |  | 134 (21%) |
| A-level or lower/Other |  | 513 (79%) |
| **Being a parent (21y)** | 9 (1.0%) |  |
| No |  | 779 (92%) |
| Yes |  | 70 (8.2%) |
| **Neighbourhood deprivation (21y)** | 23 (2.7%) |  |
| Least deprived quintile |  | 293 (35%) |
| More deprived quintiles |  | 542 (65%) |
| ***Physical and mental health*** |  |  |
| **BMI<sup>3</sup> (18y)** | 253 (29%) |  |
| <25 |  | 470 (78%) |
| >=25 |  | 135 (22%) |
| **Exercise frequency (18y)** | 375 (44%) |  |
| Weekly or more |  | 314 (65%) |
| Less than weekly |  | 169 (35%) |
| **Depressive symptoms (21y)** | 30 (3.5%) |  |
| <12 SMFQ<sup>4</sup> score |  | 653 (79%) |
| >=12 SFMQ score |  | 175 (21%) |
| <sup>1</sup> n (%); <sup>2</sup> y = approximate years of age; <sup>3</sup> BMI = Body Mass Index; <sup>4</sup> SMFQ = Short Mood and Feelings Questionnaire |  |  |

Supplementary Table S2. shows differences in characteristics by the four early-life confounders. There were differences in BMI, exercise, binge drinking and parenthood by sex and household income.

There were also sex differences in terms of other substance use and depressive symptoms. Neighbourhood deprivation differed by household income and parental smoking, and smoking frequency differed by parental smoking. Parental smoking and household income were also correlated.

### Associations of characteristics with first transitions from smoking

Figure 2 and Table 2 show the pooled results related to the 11 main characteristics of interest after adjustment for early-life confounders and weighting for selection via smoking. These are discussed below.

**Figure 2.**
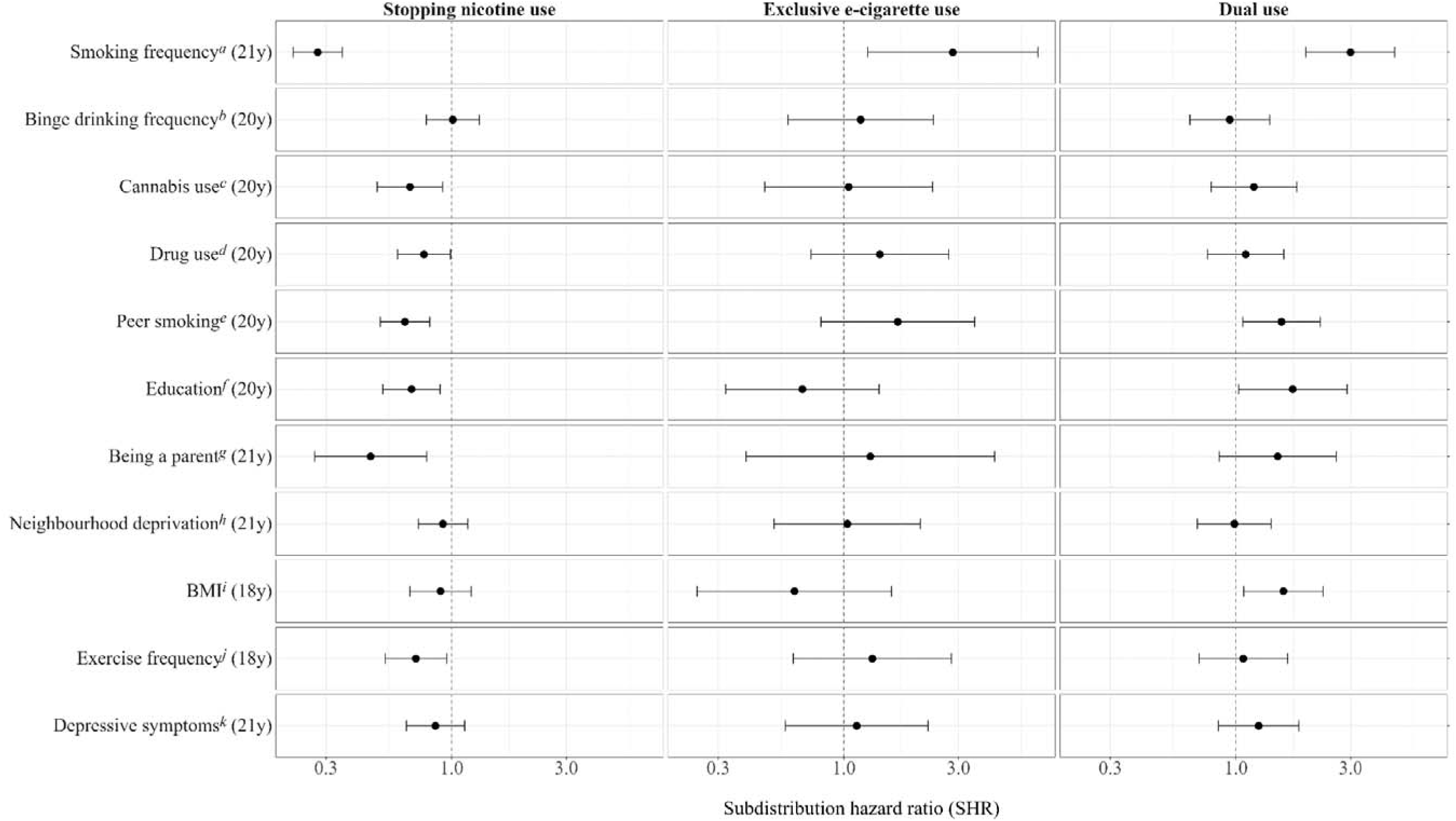
Pooled results from subdistribution discrete-time survival analyses, adjusted for sex, ethnicity, household income at 11 years [y] and parental smoking (12y), and weighted for selection via smoking. ***^a^*** occasionally smokes vs weekly or more; ***^b^*** never drinks more than 6 units, drinks monthly, or less vs weekly or more; ***^c^*** never used cannabis, not in the past year, less than monthly vs monthly or more; ***^d^*** never used drugs or not in past year vs used in past year; ***^e^*** no friends smoke, a few, or some vs most or all; ***^f^*** degree-level vs A-level, below, or other qualification; ***^g^*** not a biological or non-biological parent vs is a parent; ***^h^*** least deprived Townsend deprivation score quintile vs more deprived quintiles; ***^i^*** BMI less than 25 vs 25 or more; ***^j^*** exercised weekly or more in past year vs less than weekly; ***^k^*** Short Mood and Feelings Questionnaire (SMFQ) score of less than 12 vs score of 12 or more.

**Table 2.** Pooled summary statistics from subdistribution discrete-time survival analyses, adjusted for early-life confounders, weighted for selection via smoking.

| Characteristic | Stopping nicotine use (52%) |  |  | Exclusive e-cig use (9%) |  |  | Dual use (27%) |  |  |
| --- | --- | --- | --- | --- | --- | --- | --- | --- | --- |
|  | SHR<sup>1</sup> | 95% CI<sup>1</sup> | p | SHR<sup>1</sup> | 95% CI<sup>1</sup> | p | SHR<sup>1</sup> | 95% CI<sup>1</sup> | p |
| **Substance use** |  |  |  |  |  |  |  |  |  |
| **Smoking frequency (21y)** |  |  |  |  |  |  |  |  |  |
| Occasional | — | — |  | — | — |  | — | — |  |
| Weekly or more | 0.28 | 0.22, 0.35 | <0.001 | 2.84 | 1.26, 6.42 | 0.012 | 3.00 | 1.96, 4.59 | <0.001 |
| **Binge drinking frequency (20y)** |  |  |  |  |  |  |  |  |  |
| Never, monthly, or less | — | — |  | — | — |  | — | — |  |
| Weekly or more | 1.01 | 0.78, 1.30 | >0.9 | 1.17 | 0.58, 2.36 | 0.700 | 0.94 | 0.64, 1.38 | 0.800 |
| **Cannabis use (20y)** |  |  |  |  |  |  |  |  |  |
| Never, less than monthly, not past year | — | — |  | — | — |  | — | — |  |
| Monthly or more | 0.67 | 0.49, 0.92 | 0.013 | 1.05 | 0.47, 2.34 | >0.9 | 1.19 | 0.79, 1.79 | 0.400 |
| **Drug use (20y)** |  |  |  |  |  |  |  |  |  |
| No | — | — |  | — | — |  | — | — |  |
| Yes | 0.77 | 0.59, 0.99 | 0.042 | 1.41 | 0.73, 2.73 | 0.300 | 1.10 | 0.76, 1.58 | 0.600 |
| **Social and sociodemographic factors** |  |  |  |  |  |  |  |  |  |
| **Peer smoking (20y)** |  |  |  |  |  |  |  |  |  |
| None, a few, or some | — | — |  | — | — |  | — | — |  |
| Most or all | 0.64 | 0.50, 0.81 | <0.001 | 1.68 | 0.80, 3.50 | 0.200 | 1.55 | 1.07, 2.24 | 0.021 |
| **Education (20y)** |  |  |  |  |  |  |  |  |  |
| Degree-level | — | — |  | — | — |  | — | — |  |
| A-level or lower/Other | 0.68 | 0.52, 0.90 | 0.006 | 0.67 | 0.32, 1.40 | 0.300 | 1.72 | 1.03, 2.90 | 0.040 |
| **Being a parent (21y)** |  |  |  |  |  |  |  |  |  |
| No | — | — |  | — | — |  | — | — |  |
| Yes | 0.46 | 0.27, 0.79 | 0.005 | 1.29 | 0.39, 4.24 | 0.700 | 1.49 | 0.85, 2.62 | 0.200 |
| **Neighbourhood deprivation (21y)** |  |  |  |  |  |  |  |  |  |
| Least deprived quintile | — | — |  | — | — |  | — | — |  |
| More deprived quintiles | 0.92 | 0.73, 1.17 | 0.500 | 1.03 | 0.51, 2.08 | >0.9 | 0.98 | 0.69, 1.40 | >0.9 |
| **Physical and mental health** |  |  |  |  |  |  |  |  |  |
| **BMI (18y)** |  |  |  |  |  |  |  |  |  |
| <25 | — | — |  | — | — |  | — | — |  |
| ≥25 | 0.90 | 0.67, 1.21 | 0.500 | 0.62 | 0.25, 1.58 | 0.300 | 1.58 | 1.08, 2.31 | 0.019 |
| **Exercise frequency (18y)** |  |  |  |  |  |  |  |  |  |
| Weekly or more | — | — |  | — | — |  | — | — |  |
| Less than weekly | 0.71 | 0.53, 0.95 | 0.023 | 1.31 | 0.62, 2.80 | 0.500 | 1.07 | 0.70, 1.64 | 0.700 |
| **Depressive symptoms (21y)** |  |  |  |  |  |  |  |  |  |
| <12 SMFQ<sup>2</sup> score | — | — |  | — | — |  | — | — |  |
| ≥12 SMFQ score | 0.86 | 0.65, 1.13 | 0.300 | 1.13 | 0.57, 2.24 | 0.700 | 1.24 | 0.85, 1.83 | 0.300 |

<sup>1</sup> SHR = Subdistribution Hazard Ratio, CI = Confidence Interval; <sup>2</sup> SMFQ = Short Mood and Feelings Questionnaire

#### Substance use

Smoking frequency at age 21 was the only characteristic associated with all possible transitions from smoking. Participants who smoked weekly or more were less likely to transition to stopping nicotine use (SHR = 0.28, 95% CI = 0.22-0.35), and more likely to transition to exclusive e-cigarette use (SHR = 2.84, 95% CI = 1.26-6.42) and dual use (SHR = 3.00, 95% CI = 1.96-4.59), at each time-point.

There was no evidence to suggest binge drinking was associated with any transitions from smoking in this study. Participants who, at age 20, reported using cannabis monthly or more in the past year (SHR = 0.67, 95% CI = 0.49-0.92), or using other drugs in the past year (SHR = 0.77, 95% CI = 0.59-0.99) were less likely to transition to stopping nicotine use. There was little to no evidence that other substance use was associated with transitions to using e-cigarettes.

#### Social and sociodemographic factors

Participants who reported that most or all their friends smoked were less likely to transition to stopping nicotine use (SHR = 0.64, 95% CI = 0.50-0.81), and more likely to transition to dual use (SHR = 1.55, 95% CI = 1.07-2.24).

Participants who reported having fewer educational qualifications at age 20 were also less likely to transition to stopping nicotine use (SHR = 0.68, 95% CI = 0.52-0.90) and more likely to transition to dual use (SHR = 1.72, 95% CI = 1.03-2.90).

There was weak evidence to suggest peer smoking and education was associated with transitions to exclusive e-cigarette use given the large amount of uncertainty reflected in the wide confidence intervals. However the point estimate related to the association of peer smoking with exclusive e-cigarette use was similar as for dual use. Conversely the point estimate reflecting the association of education with exclusive e-cigarette use showed the opposite direction of association compared to dual use.

Participants who reported being a parent at age 21 were less likely to transition to stopping nicotine use (SHR = 0.46, 95% CI = 0.27-0.79). There was little to no evidence that parenthood was associated with transitions to using e-cigarettes.

There was no clear evidence to suggest neighbourhood deprivation at age 21 was associated with any transitions from smoking in this study.

#### Physical and mental health

Participants with a BMI over or equal to 25 at age 18 were more likely to transition to dual use (SHR = 1.58, 95% CI = 1.08-2.31). There was little to no evidence that BMI was associated with transitions to stopping nicotine use, or to exclusive e-cigarette use.

Participants who exercised less than weekly in the past year at age 18 were less likely to transition to stopping nicotine use (SHR = 0.71, 95% CI = 0.53-0.95). There was little to no evidence that exercise frequency was associated with transitions to using e-cigarettes.

There was little to no evidence to suggest that depressive symptoms at age 21 was associated with any transitions from smoking in this study.

#### Other analyses

Results related to the four early-life confounders are shown in Supplementary Table S3 but not discussed here as these coefficients cannot be interpreted as the effect of each on transitions from smoking due to mutual adjustment.

Results from unweighted, unadjusted, and complete case analyses are shown in Supplementary Tables S4-6. Results from unweighted and unadjusted analyses were similar to those described above, while the unweighted complete case analyses showed some differences. When analyses were limited to complete cases there was no association of parenthood at age 21 with transitions to stopping nicotine use, but parents did appear more likely to transition to dual use. Unweighted complete case analyses also showed no association of smoking frequency with transitions to exclusive e-cigarette use, nor BMI and peer smoking with transitions to dual use. However these results are likely more biased than those described above due to missing data.

## Discussion

In a sample of young adults who smoked tobacco, this study explored predictors of first reported transitions from smoking (21y) to stopping nicotine use, exclusive e-cigarette use, and dual use, in early adulthood (22-30y) within a UK setting. More participants transitioned to stopping nicotine use than to using e-cigarettes, and transitions to dual use were more common than to exclusive e-cigarette use. Participants who exercised infrequently at age 18, used cannabis or other drugs at age 20, or who were parents at age 21, had a decreased likelihood of stopping nicotine use but were not more (or less) likely to use e-cigarettes. Participants who had lower education or reported greater peer smoking at age 20, or who frequently used tobacco at age 21, also had a decreased likelihood of stopping nicotine use, and were more likely to progress to dual use of tobacco and e-cigarettes. Smoking frequency was also associated with switching to exclusively using e-cigarettes. BMI was associated with transition to dual use only. No effect of binge drinking at age 20, nor neighbourhood deprivation or depressive symptoms at age 21, on transitions from smoking was detected.

There are many barriers to stopping nicotine use. The groups we have identified who are less likely to stop using nicotine show how complex lifestyle behaviours and stressors may make quitting more difficult. However despite the increased prevalence of e-cigarette use amongst those who previously or currently smoked^1^ these groups appear to vary in terms of their e-cigarette use. We identified several groups who may not take up using e-cigarettes. Cannabis and tobacco are linked due to their co-use in the UK^41,42^ but previous findings relating to cannabis and e-cigarette use have been mixed^43,44^. Higher levels of physical activity have been linked to higher e-cigarette use in young adults^45^, but lower cigarette smoking^46^. Early parenthood has been linked to persistent smoking^47^ but parents who dual use may contemplate smoking cessation more than parents who only smoke cigarettes^48^. Crucially, the continued use of tobacco in these groups is likely to exacerbate existing health inequalities as those using cannabis are at increased risk of mental health disorders^49,50^, the combined use of tobacco and lower physical activity has additive impacts on poor physical health^51^, and continued use of tobacco amongst young parents makes the next generation more likely to smoke themselves^52^. Consequently, these groups represent key targets for smoking cessation and harm reduction interventions.

Several groups were identified who were more likely to transition to dual use of tobacco and e-cigarettes. Findings are mixed regarding harm reduction via dual use. There is evidence that dual use is related to reduced tobacco cigarette consumption^53^, but other evidence indicates that dual use may not provide substantial harm reduction^16,17,54^. Individuals who have higher BMI^55^, higher education^56^, more social connections who smoke^57^, and smoke more cigarettes themselves^58,59^, have previously been identified as more likely to use e-cigarettes. It is unknown whether these individuals have transitioned to dual use with an aim of ceasing smoking as previously reported motivations for dual use focus on pleasure, rather than desire to quit^15^. Dual use may then reflect ambivalence toward quitting or an attempt to mitigate harm while maintaining nicotine dependence. Nevertheless, our findings suggest that dual use may be particularly common among people with more entrenched smoking habits or social influences, who may in turn not feel ready or able to fully quit smoking. Understanding how these groups can be encouraged to solely use e-cigarettes, which has lower impacts on health^54^, is then an important step for public health. Our findings suggest that while smoking frequency and peer smoking may influence e-cigarette use generally, education and BMI may play distinct roles in influencing whether individuals exclusively use e-cigarettes or engage in dual use. This supports previous work showing that e-cigarette use often co-exists with tobacco use within lower socioeconomic groups rather than displacing it^60,61^.

A lot of research into e-cigarette use focusses on individuals who previously did not smoke^57,62^, whereas our focus on individuals who had already begun tobacco use fulfils important gaps in the literature, and offers insights and implications for tobacco cessation and harm reduction. Current intervention to tobacco cessation focusses on a variety of strategies such as behavioural therapies^63^, pharmacological treatments^64^, financial incentives^65^. and innovative digital approaches^66–69^. Our present work suggests that these efforts could benefit from being better tailored towards the groups indicated. Tailored programmes that help people who smoke switch to less harmful alternatives to smoking, and help people who dual use switch completely to e-cigarettes, could be developed and better promoted towards people who are more demographically vulnerable or dependant on cigarettes^70^. However currently the effects of tailoring smoking cessation interventions for disadvantaged people has been limited^71^, and programs that aim to integrate and address substance use, physical inactivity and smoking concurrently need to be improved^72^.

E-cigarettes are providing a new avenue to help people quit smoking, where behavioural support combined with e-cigarette use has been shown to be particularly effective^73^ but it is important to ensure any ensuing reductions in smoking-related harms are equitable^74^. In the UK existing NHS smoking cessation services have achieved great success but whether these interventions have a differential impact on particular subpopulations is less clear^75^. The recent NHS “Swap to Stop” scheme operates under a harm-reduction framework rather than a clinical intervention. Our findings underscore the importance of understanding e-cigarettes within a harm-reduction context and may help improve the efficacy of public health initiatives like this that encourage people who smoke to switch to vaping.

### Strengths and limitations

The strengths of this study include the length of follow up data spanning young adulthood, the use of a phenotypically rich cohort with many measures of relevant risk factors, the use of a competing risk framework, and the use of multiple imputation and weights to mitigate complete case and selection bias respectively to garner more robust findings.

Limitations include the small sample size, measurement error where complex characteristics have been simplified, and potential confounding or interactions between the characteristics being separately investigated. Left and interval censoring is another issue that may lead to less precise estimates.

Current e-cigarette use prior to 22 years was not known, nor the exact timings of reported transitions. Given periods of e-cigarette use may be shorter than periods of non-use, and that there were large gaps between measurements of nicotine use, investigating first transitions from smoking using interval-censored data may be better equipped to detect abstinence from nicotine rather than e-cigarette use. Other sources of nicotine such as NRT were not investigated. Findings may also be subject to attrition bias as continued participation in ALSPAC has been shown to be non-random^76,77^ and our sample is limited to those who participated in the 21+ questionnaire.

The landscape of e-cigarettes in the UK is rapidly evolving and increasing in popularity^78^ where the use of e-cigarettes within England among smokers and recent ex-smokers plateaued from 2013 to 2020 but has grown since^79^. This makes it difficult to understand the influence of participant characteristics independent of their current technological, social and political contexts. In this cohort stopping nicotine use often occurred in the earliest timepoints before e-cigarettes were as popular or as effective at delivering nicotine as they are today^6,80^. Here participants who were able to quit smoking unaided or with the use of traditional NRT did so, while many participants who did not quit still later engaged with e-cigarettes. This may explain the larger proportion of transitions to dual use compared to exclusive e-cigarette use.

Larger and more well powered studies will be needed to investigate how characteristics such as depressive symptoms or neighbourhood deprivation, that may contribute smaller effects and vary over time, influence transitions from smoking. Such studies could also be used to explore subsequent transitions between nicotine use states, e.g. stopping nicotine use after using e-cigarettes, via multi-state model analysis. Future work should also examine the complex interplay between different characteristics and motivations on quitting smoking or using e-cigarettes.

### Implications

We identified many groups who likely need greater efforts from stop smoking interventions. Some of these groups do appear to be using e-cigarettes, but often by combining e-cigarette use with tobacco and so may not fully benefit from their harm reduction potential. Other groups may not be engaging with e-cigarettes, or find it more difficult than others to feel encouraged to switch from smoking to vaping. Future policies and interventions should address the potential harms of dual use versus completely switching to e-cigarettes, and encourage safer alternative sources of nicotine, especially if they are unable or unwilling to quit smoking unaided or with other nicotine replacement methods.

Together these insights could be used to improve the effectiveness of current tobacco harm reduction interventions but careful consideration of policies around e-cigarettes is needed to ensure existing health inequalities are not further perpetuated.

## Data Access and Sharing

Data used in this project and any resulting data from the analyses are available on request to the ALSPAC Executive Committee and subject to a data access fee. Ethical approval for the study was obtained from the ALSPAC Law and Ethics Committee and Local Research Ethics Committees (NHS Haydock REC: 10/H1010/70). Informed consent for the use of data collected via questionnaires and clinics was obtained from participants following the recommendations of the ALSPAC Ethics and Law Committee at the time. Consent for biological samples has been collected in accordance with the Human Tissue Act (2004). Data access for this project was granted (B3499/B4347) prior to this study. Datasets were created using syntax templates from the 25^th^ January 2024.

The proposed analysis of data was pre-registered on the Open Science Framework (OSF) here: https://osf.io/nuz5b. Deviations from this pre-registration are described in Supplementary Text S1. The code used for data analysis is available in a github repository here: https://github.com/alexandrayas/ALSPAC_CRUK_smkvap/tree/main/Transitions%20from%20smoking.

## Author contribution

**AA**: conceptualization; formal analysis; visualization; writing – original draft preparation; writing – review & editing. **JH**: conceptualization; formal analysis; supervision; writing – review & editing. **JK**: conceptualization; formal analysis; writing – review & editing. **HJ**: conceptualization; formal analysis; writing – review & editing. **HS**: funding acquisition; conceptualization; supervision; writing – review & editing. **MM**: funding acquisition; writing – review & editing. **LH**: conceptualization; formal analysis; supervision; writing – review & editing. **EC**: conceptualization; formal analysis; visualization; supervision; writing – review & editing.

## Data Availability

Data used in this project are available on request to the ALSPAC Executive Committee and subject to a data access fee.

https://www.bristol.ac.uk/alspac/researchers/access/

## Acknowledgements

We are extremely grateful to all the families who took part in this study, the midwives for their help in recruiting them, and the whole ALSPAC team, which includes interviewers, computer and laboratory technicians, clerical workers, research scientists, volunteers, managers, receptionists, and nurses.

## References

1. ASH. Smoking Statistics. ASH. October 2023. Accessed July 1, 2024. https://ash.org.uk/resources/view/smoking-statistics

2. Goniewicz ML, Knysak J, Gawron M, et al. Levels of selected carcinogens and toxicants in vapour from electronic cigarettes. Tob Control. 2014;23(2):133–139. doi:10.1136/tobaccocontrol-2012-050859

3. Hartmann-Boyce J, McRobbie H, Lindson N, et al. Electronic cigarettes for smoking cessation. Cochrane Database Syst Rev. 2020;(10). doi:10.1002/14651858.CD010216.pub4

4. Hartmann-Boyce J, Lindson N, Butler AR, et al. Electronic cigarettes for smoking cessation. Cochrane Database Syst Rev. 2022;11(11):CD010216. doi:10.1002/14651858.CD010216.pub7

5. PHE. Electronic cigarettes: report commissioned by PHE. Published online 2014.

6. ASH. Use of e-cigarettes among adults in Great Britain. ASH. August 2023. Accessed June 28, 2024. https://ash.org.uk/resources/view/use-of-e-cigarettes-among-adults-in-great-britain-2021

7. Beard E, West R, Michie S, Brown J. Association of prevalence of electronic cigarette use with smoking cessation and cigarette consumption in England: a time-series analysis between 2006 and 2017. Addict Abingdon Engl. 2020;115(5):961–974. doi:10.1111/add.14851

8. Beard E, Brown J, Shahab L. Association of quarterly prevalence of e-cigarette use with ever regular smoking among young adults in England: a time-series analysis between 2007 and 2018. Addict Abingdon Engl. 2022;117(8):2283–2293. doi:10.1111/add.15838

9. McNeill A, Brose L, Calder R, Bauld L, Robson D. Vaping in England: an evidence update including mental health. Published online 2020.

10. McNeill A, Brose L, Calder R, Simonavicius E, Robson D. Vaping in England: 2021 evidence update summary. GOV.UK. February 23, 2021. Accessed March 18, 2024. https://www.gov.uk/government/publications/vaping-in-england-evidence-update-february-2021/vaping-in-england-2021-evidence-update-summary

11. MHRA. Nicotine replacement therapy and harm reduction. GOV.UK. November 12, 2014. Accessed March 18, 2024. https://www.gov.uk/drug-safety-update/nicotine-replacement-therapy-and-harm-reduction

12. TPD. Directive 2014/40/EU of the European Parliament and of the Council of 3 April 2014 on the approximation of the laws, regulations and administrative provisions of the Member States concerning the manufacture, presentation and sale of tobacco and related products and repealing Directive 2001/37/EC (Text with EEA relevance). https://webarchive.nationalarchives.gov.uk/eu-exit/https://eur-lex.europa.eu/legal-content/EN/TXT/?uri=CELEX:02014L0040-20150106. 2014. Accessed March 18, 2024. https://www.legislation.gov.uk/eudr/2014/40/title/II

13. Rough E. The regulation of e-cigarettes. Published online January 7, 2024. Accessed July 1, 2024. https://commonslibrary.parliament.uk/research-briefings/cbp-8114/

14. DHSC. Smokers urged to swap cigarettes for vapes in world first scheme. GOV.UK. Accessed July 18, 2024. https://www.gov.uk/government/news/smokers-urged-to-swap-cigarettes-for-vapes-in-world-first-scheme

15. Temourian AA, Song AV, Halliday DM, Gonzalez M, Epperson AE. Why do smokers use e-cigarettes? A study on reasons among dual users. Prev Med Rep. 2022;29:101924. doi:10.1016/j.pmedr.2022.101924

16. Shahab L, Goniewicz ML, Blount BC, et al. Nicotine, Carcinogen, and Toxin Exposure in Long-Term E-Cigarette and Nicotine Replacement Therapy Users: A Cross-sectional Study. Ann Intern Med. 2017;166(6):390–400. doi:10.7326/M16-1107

17. Stokes AC, Xie W, Wilson AE, et al. Association of Cigarette and Electronic Cigarette Use Patterns with Levels of Inflammatory and Oxidative Stress Biomarkers among US Adults, Population Assessment of Tobacco and Health Study. Circulation. 2021;143(8):869–871. doi:10.1161/CIRCULATIONAHA.120.051551

18. Snell LM, Barnes AJ, Nicksic NE. A Longitudinal Analysis of Nicotine Dependence and Transitions From Dual Use of Cigarettes and Electronic Cigarettes: Evidence From Waves 1–3 of the PATH Study. J Stud Alcohol Drugs. 2020;81(5):595–603. doi:10.15288/jsad.2020.81.595

19. Conner M, Grogan S, Simms-Ellis R, et al. Patterns and predictors of e-cigarette, cigarette and dual use uptake in UK adolescents: evidence from a 24-month prospective study. Addiction. 2019;114(11):2048–2055. doi:10.1111/add.14723

20. Kalan ME, Brewer NT. Longitudinal transitions in e-cigarette and cigarette use among US adults: prospective cohort study. Lancet Reg Health – Am. 2023;22. doi:10.1016/j.lana.2023.100508

21. Kinnunen JM, Ollila H, Minkkinen J, Lindfors PL, Rimpelä AH. A Longitudinal Study of Predictors for Adolescent Electronic Cigarette Experimentation and Comparison with Conventional Smoking. Int J Environ Res Public Health. 2018;15(2):305. doi:10.3390/ijerph15020305

22. Tokle R, Brunborg GS, Vedøy TF. Adolescents’ Use of Nicotine-Free and Nicotine E-Cigarettes: A Longitudinal Study of Vaping Transitions and Vaper Characteristics. Nicotine Tob Res. 2022;24(3):400–407. doi:10.1093/ntr/ntab192

23. Niaura R, Rich I, Johnson AL, et al. Young Adult Tobacco and E-cigarette Use Transitions: Examining Stability Using Multistate Modeling. Nicotine Tob Res. 2020;22(5):647–654. doi:10.1093/ntr/ntz030

24. Brouwer AF, Jeon J, Hirschtick JL, et al. Transitions between cigarette, ENDS and dual use in adults in the PATH study (waves 1–4): multistate transition modelling accounting for complex survey design. Tob Control. 2022;31(3):424–431. doi:10.1136/tobaccocontrol-2020-055967

25. Parnham JC, Vrinten C, Radó MK, Bottle A, Filippidis FT, Laverty AA. Multistate transition modelling of e-cigarette use and cigarette smoking among youth in the UK. Tob Control. 2024;33(4):489–496. doi:10.1136/tc-2022-057777

26. Nam JK, Piper ME, Tong Z, et al. Dependence motives and use contexts that predicted smoking cessation and vaping cessation: A two-year longitudinal study with 13 waves. Drug Alcohol Depend. 2023;250:110871. doi:10.1016/j.drugalcdep.2023.110871

27. Boyd A, Golding J, Macleod J, et al. Cohort Profile: The ‘Children of the 90s’—the index offspring of the Avon Longitudinal Study of Parents and Children. Int J Epidemiol. 2013;42(1):111–127. doi:10.1093/ije/dys064

28. Fraser A, Macdonald-Wallis C, Tilling K, et al. Cohort Profile: The Avon Longitudinal Study of Parents and Children: ALSPAC mothers cohort. Int J Epidemiol. 2013;42(1):97–110. doi:10.1093/ije/dys066

29. Northstone K, Lewcock M, Groom A, et al. The Avon Longitudinal Study of Parents and Children (ALSPAC): an update on the enrolled sample of index children in 2019. Wellcome Open Res. 2019;4:51. doi:10.12688/wellcomeopenres.15132.1

30. Major-Smith D, Heron J, Fraser A, Lawlor DA, Golding J, Northstone K. The Avon Longitudinal Study of Parents and Children (ALSPAC): a 2022 update on the enrolled sample of mothers and the associated baseline data. Wellcome Open Res. 2022;7:283. doi:10.12688/wellcomeopenres.18564.1

31. Northstone K, Ben-Shlomo Y, Teyhan A, et al. The Avon Longitudinal Study of Parents and children ALSPAC G0…Wellcome Open Res. Published online July 2023. doi:10.12688/wellcomeopenres.18782.2

32. Harris PA, Taylor R, Thielke R, Payne J, Gonzalez N, Conde JG. Research electronic data capture (REDCap)--a metadata-driven methodology and workflow process for providing translational research informatics support. J Biomed Inform. 2009;42(2):377–381. doi:10.1016/j.jbi.2008.08.010

33. StataCorp. Stata Statistical Software: Release 16. Published online 2019.

34. R Core Team. R: A Language and Environment for Statistical Computing. Published online 2023. www.R-project.org

35. Berger M, Schmid M, Welchowski T, Schmitz-Valckenberg S, Beyersmann J. Subdistribution hazard models for competing risks in discrete time. Biostatistics. 2020;21(3):449–466. doi:10.1093/biostatistics/kxy069

36. Austin PC, Fine JP. Practical recommendations for reporting FinelGray model analyses for competing risk data. Stat Med. 2017;36(27):4391–4400. doi:10.1002/sim.7501

37. van Buuren S. Multiple imputation of discrete and continuous data by fully conditional specification. Stat Methods Med Res. 2007;16(3):219–242. doi:10.1177/0962280206074463

38. van Buuren S, Groothuis-Oudshoorn K. mice: Multivariate Imputation by Chained Equations in R. J Stat Softw. 2011;45(3):1–67. doi:10.18637/jss.v045.i03

39. Digitale JC, Martin JN, Glidden DV, Glymour MM. Key concepts in clinical epidemiology: collider-conditioning bias. J Clin Epidemiol. 2023;161:152–156. doi:10.1016/j.jclinepi.2023.07.004

40. Tattan-Birch H, Marsden J, West R, Gage SH. Assessing and addressing collider bias in addiction research: the curious case of smoking and COVID-19. Addiction. 2021;116(5):982–984. doi:10.1111/add.15348

41. Hindocha C, Freeman TP, Ferris JA, Lynskey MT, Winstock AR. No smoke without tobacco: A global overview of cannabis and tobacco routes of administration and their association with intention to quit. Front Psychiatry. 2016;7.

42. Hindocha C, Brose LS, Walsh H, Cheeseman H. Cannabis use and co-use in tobacco smokers and non-smokers: prevalence and associations with mental health in a cross-sectional, nationally representative sample of adults in Great Britain, 2020. Addiction. 2021;116(8):2209-2219. doi:10.1111/add.15381

43. Gravely S, Driezen P, McClure EA, et al. Differences between adults who smoke cigarettes daily and do and do not co-use cannabis: Findings from the 2020 ITC four country smoking and vaping survey. Addict Behav. 2022;135:107434. doi:10.1016/j.addbeh.2022.107434

44. Rice M, Nollen NL, Ahluwalia JS, Benowitz N, Woodcock A, Pulvers K. Effects of Marijuana Use on Smokers Switching to E-Cigarettes in a Randomized Clinical Trial. Nicotine Tob Res. 2022;24(7):994–1002. doi:10.1093/ntr/ntac008

45. Pokhrel P, Schmid S, Pagano I. Physical Activity and Use of Cigarettes and E-Cigarettes Among Young Adults. Am J Prev Med. 2020;58(4):580–583. doi:10.1016/j.amepre.2019.10.015

46. Kujala UM, Kaprio J, Rose RJ. Physical activity in adolescence and smoking in young adulthood: a prospective twin cohort study. Addict Abingdon Engl. 2007;102(7):1151–1157. doi:10.1111/j.1360-0443.2007.01858.x

47. Jefferis BJMH, Power C, Graham H, Manor O. Effects of Childhood Socioeconomic Circumstances on Persistent Smoking. Am J Public Health. 2004;94(2):279–285.

48. Nabi-Burza E, Regan S, Walters BH, et al. Parental Dual Use of e-Cigarettes and Traditional Cigarettes. Acad Pediatr. 2019;19(7):842–848. doi:10.1016/j.acap.2019.04.001

49. Moore TH, Zammit S, Lingford-Hughes A, et al. Cannabis use and risk of psychotic or affective mental health outcomes: a systematic review. The Lancet. 2007;370(9584):319-328. doi:10.1016/S0140-6736(07)61162-3

50. Gobbi G, Atkin T, Zytynski T, et al. Association of Cannabis Use in Adolescence and Risk of Depression, Anxiety, and Suicidality in Young Adulthood. JAMA Psychiatry. 2019;76(4):426–434. doi:10.1001/jamapsychiatry.2018.4500

51. Jackson SE, Brown J, Ussher M, Shahab L, Steptoe A, Smith L. Combined health risks of cigarette smoking and low levels of physical activity: a prospective cohort study in England with 12-year follow-up. BMJ Open. 2019;9(11):e032852. doi:10.1136/bmjopen-2019-032852

52. Alves J, Perelman J, Ramos E, Kunst AE. Intergenerational transmission of parental smoking: when are offspring most vulnerable? Eur J Public Health. 2022;32(5):741–746. doi:10.1093/eurpub/ckac065

53. Jackson SE, Farrow E, Brown J, Shahab L. Is dual use of nicotine products and cigarettes associated with smoking reduction and cessation behaviours? A prospective study in England. BMJ Open. 2020;10(3):e036055. doi:10.1136/bmjopen-2019-036055

54. Glantz SA, Nguyen N, Oliveira da Silva AL. Population-Based Disease Odds for E-Cigarettes and Dual Use versus Cigarettes. NEJM Evid. 2024;3(3):EVIDoa2300229. doi:10.1056/EVIDoa2300229

55. Lanza HI, Pittman P, Batshoun J. Obesity and Cigarette Smoking: Extending the Link to E cigarette/Vaping Use. Am J Health Behav. 2017;41(3):338–347. doi:10.5993/AJHB.41.3.13

56. Wilson FA, Wang Y. Recent Findings on the Prevalence of E-Cigarette Use Among Adults in the U.S. Am J Prev Med. 2017;52(3):385–390. doi:10.1016/j.amepre.2016.10.029

57. Amin S, Dunn AG, Laranjo L. Social Influence in the Uptake and Use of Electronic Cigarettes: A Systematic Review. Am J Prev Med. 2020;58(1):129–141. doi:10.1016/j.amepre.2019.08.023

58. Brown J, West R, Beard E, Michie S, Shahab L, McNeill A. Prevalence and characteristics of e-cigarette users in Great Britain: Findings from a general population survey of smokers. Addict Behav. 2014;39(6):1120–1125. doi:10.1016/j.addbeh.2014.03.009

59. Pearson JL, Stanton CA, Cha S, Niaura RS, Luta G, Graham AL. E-Cigarettes and Smoking Cessation: Insights and Cautions From a Secondary Analysis of Data From a Study of Online Treatment-Seeking Smokers. Nicotine Tob Res. 2015;17(10):1219–1227. doi:10.1093/ntr/ntu269

60. Thirlway F. How will e-cigarettes affect health inequalities? Applying Bourdieu to smoking and cessation. Int J Drug Policy. 2018;54:99–104. doi:10.1016/j.drugpo.2018.01.009

61. Lucherini M, Hill S, Smith K. Potential for non-combustible nicotine products to reduce socioeconomic inequalities in smoking: a systematic review and synthesis of best available evidence. BMC Public Health. 2019;19(1):1469. doi:10.1186/s12889-019-7836-4

62. Lieu TA, Bibbins-Domingo K. E-Cigarette Use in Adolescents and Adults—A JAMA Collection. JAMA. 2024;332(9):709–710. doi:10.1001/jama.2024.15912

63. Hartmann-Boyce J, Livingstone-Banks J, Ordóñez-Mena JM, et al. Behavioural interventions for smoking cessation: an overview and network metalanalysis - Hartmann-Boyce, J - 2021 | Cochrane Library. Accessed October 8, 2024. https://www.cochranelibrary.com/cdsr/doi/10.1002/14651858.CD013229.pub2/full

64. Cahill K, Stevens S, Lancaster T. Pharmacological Treatments for Smoking Cessation. JAMA. 2014;311(2):193–194. doi:10.1001/jama.2013.283787

65. Giles EL, Robalino S, McColl E, Sniehotta FF, Adams J. The Effectiveness of Financial Incentives for Health Behaviour Change: Systematic Review and Meta-Analysis. PLOS ONE. 2014;9(3):e90347. doi:10.1371/journal.pone.0090347

66. Ubhi HK, Michie S, Kotz D, Wong WC, West R. A Mobile App to Aid Smoking Cessation: Preliminary Evaluation of SmokeFree28. J Med Internet Res. 2015;17(1):e3479. doi:10.2196/jmir.3479

67. Scott-Sheldon LAJ, Lantini RC, Jennings EG, et al. Text Messaging-Based Interventions for Smoking Cessation: A Systematic Review and Meta-Analysis. JMIR MHealth UHealth. 2016;4(2):e5436. doi:10.2196/mhealth.5436

68. Regmi K, Kassim N, Ahmad N, Tuah NA. Effectiveness of Mobile Apps for Smoking Cessation: A Review. Tob Prev Cessat. 2017;3:12. doi:10.18332/tpc/70088

69. Eghdami S, Ahmadkhaniha HR, Baradaran HR, Hirbod-Mobarakeh A. Ecological momentary interventions for smoking cessation: a systematic review and meta-analysis. Soc Psychiatry Psychiatr Epidemiol. 2023;58(10):1431–1445. doi:10.1007/s00127-023-02503-2

70. Bauld L, Judge K, Platt S. Assessing the impact of smoking cessation services on reducing health inequalities in England: observational study. Tob Control. 2007;16(6):400–404. doi:10.1136/tc.2007.021626

71. Kock L, Brown J, Hiscock R, Tattan-Birch H, Smith C, Shahab L. Individual-level behavioural smoking cessation interventions tailored for disadvantaged socioeconomic position: a systematic review and meta-regression. Lancet Public Health. 2019;4(12):e628–e644. doi:10.1016/S2468-2667(19)30220-8

72. Hovhannisyan K, Rasmussen M, Adami J, Wikström M, Tønnesen H. Evaluation of Very Integrated Program: Health Promotion for Patients With Alcohol and Drug Addiction—A Randomized Trial. Alcohol Clin Exp Res. 2020;44(7):1456–1467. doi:10.1111/acer.14364

73. Hajek P, Phillips-Waller A, Przulj D, et al. A Randomized Trial of E-Cigarettes versus Nicotine-Replacement Therapy. N Engl J Med. 2019;380(7):629–637. doi:10.1056/NEJMoa1808779

74. Buckell J, Fucito LM, Krishnan-Sarin S, O’Malley S, Sindelar JL. Harm reduction for smokers with little to no quit interest: can tobacco policies encourage switching to e-cigarettes? Tob Control. 2023;32(e2):e173–e179. doi:10.1136/tobaccocontrol-2021-057024

75. Bauld L, Bell K, McCullough L, Richardson L, Greaves L. The effectiveness of NHS smoking cessation services: a systematic review. J Public Health. 2010;32(1):71–82. doi:10.1093/pubmed/fdp074

76. Howe LD, Tilling K, Galobardes B, Lawlor DA. Loss to follow-up in cohort studies: bias in estimates of socioeconomic inequalities. Epidemiol Camb Mass. 2013;24(1):1–9. doi:10.1097/EDE.0b013e31827623b1

77. Taylor AE, Jones HJ, Sallis H, et al. Exploring the association of genetic factors with participation in the Avon Longitudinal Study of Parents and Children. Int J Epidemiol. 2018;47(4):1207–1216. doi:10.1093/ije/dyy060

78. Tattan-Birch H, Jackson SE, Kock L, Dockrell M, Brown J. Rapid growth in disposable e-cigarette vaping among young adults in Great Britain from 2021 to 2022: a repeat cross-sectional survey. Addict Abingdon Engl. 2023;118(2):382–386. doi:10.1111/add.16044

79. Buss V, Kock L, West R, Beard E, Kale D, Brown J. Trends in electronic cigarette use in England. Smoking in England. July 18, 2024. Accessed August 6, 2024. https://smokinginengland.info/graphs/e-cigarettes-latest-trends

80. Voos N, Goniewicz ML, Eissenberg T. What is the nicotine delivery profile of electronic cigarettes? Expert Opin Drug Deliv. 2019;16(11):1193–1203. doi:10.1080/17425247.2019.1665647

