## Supplementary Materials for "Transitions from smoking to exclusive e-cigarette use, dual use, or stopping nicotine use in ALSPAC and their association with modifiable and sociodemographic factors"

**Contents**

**Supplementary Text**

**Supplementary Figures**

**Supplementary Tables**

Supplementary Text S1. Divergence from pre-registration

The pre-registration for this study (<https://osf.io/u6g4s/>) stated that logistic regression and cause-specific survival analyses were to be used to investigate associations between participant characteristics and first reported transitions from smoking. However, these were not taken forward given that logistic regression does not consider the time-to-event, and cause-specific survival analysis does not consider the occurrence of competing events. As the investigated events are transient and not absorbing, sub-distribution survival analysis then better fit the research question.

The pre-registration also stated that multi-state models were to be fitted to investigate all possible transitions, not just the first reported transitions from smoking, however this was not possible due to limited sample size and statistical power.

Supplementary Figure S1. Flowchart showing attrition and available sample size

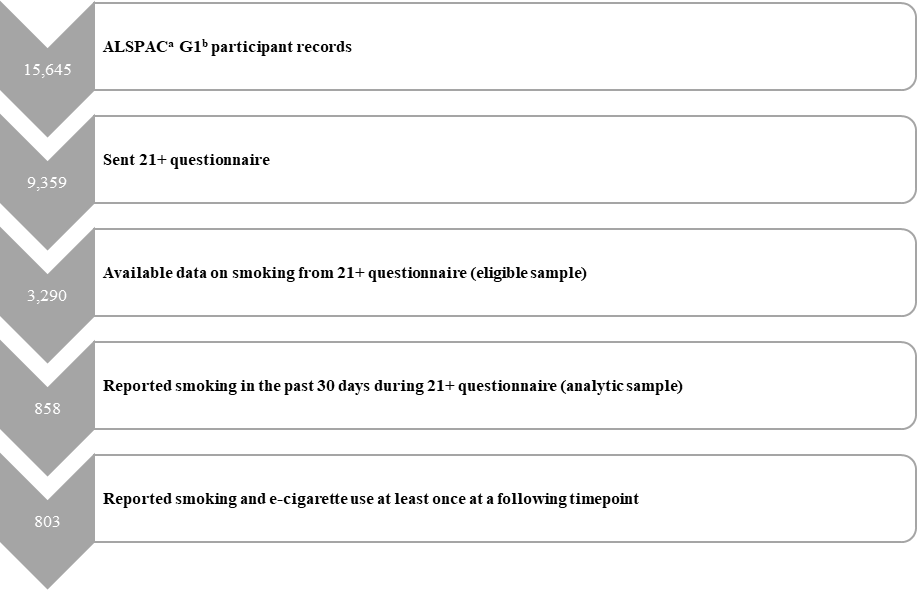

**^a^Avon Longitudinal Study of Parents and Children; ^b^ Offspring of women resident in Avon, UK with expected delivery dates between 1st April 1991 and 31st December 1992 who took part in the study**

15,645

ALSPACa G1b participant records

9,359

Sent 21+ questionnaire

3,290

Available data on smoking from 21+ questionnaire (eligible sample)

858

Reported smoking in the past 30 days during 21+ questionnaire (analytic sample)

803

Reported smoking and e-cigarette use at least once at a following timepoint

Supplementary Figure S2. Illustrative example showing how first reported transitions from smoking are identified using synthetic data for five participants

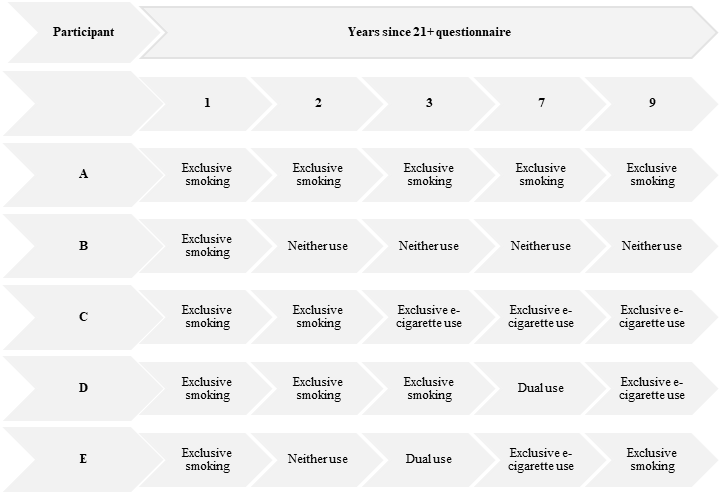

**Each row (light grey) represents synthetic data for each of five (A-E) participants. Each column (light grey) represents synthetic reported nicotine use by each participant at each of five timepoints (1, 2, 3, 7, and 9 years following the 21+ questionnaire). The dark grey boxes represent the derived first reported transition from smoking for each participant. For example, participant A only reported exclusively smoking and so did not transition. Participant D reported exclusively smoking at all timepoints up to 7 years, and then reported dual use (of cigarettes and e-cigarettes) and so their first transition from smoking would be to dual use.**

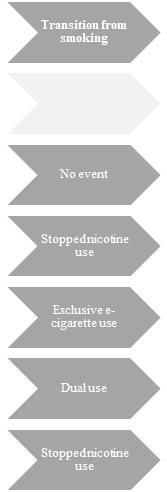

Participant

Years since 21+ questionnaire

1

2

3

7

9

A

Exclusive smoking

Exclusive smoking

Exclusive smoking

Exclusive smoking

Exclusive smoking

B

Exclusive smoking

Neither use

Neither use

Neither use

Neither use

C

Exclusive smoking

Exclusive smoking

Exclusive e-cigarette use

Exclusive e-cigarette use

Exclusive e-cigarette use

D

Exclusive smoking

Exclusive smoking

Exclusive smoking

Dual use

Exclusive e-cigarette use

E

Exclusive smoking

Neither use

Dual use

Exclusive e-cigarette use

Exclusive smoking

Transition from smoking

No event

Stopped nicotine use

Exclusive e-cigarette use

Dual use

Stopped nicotine use

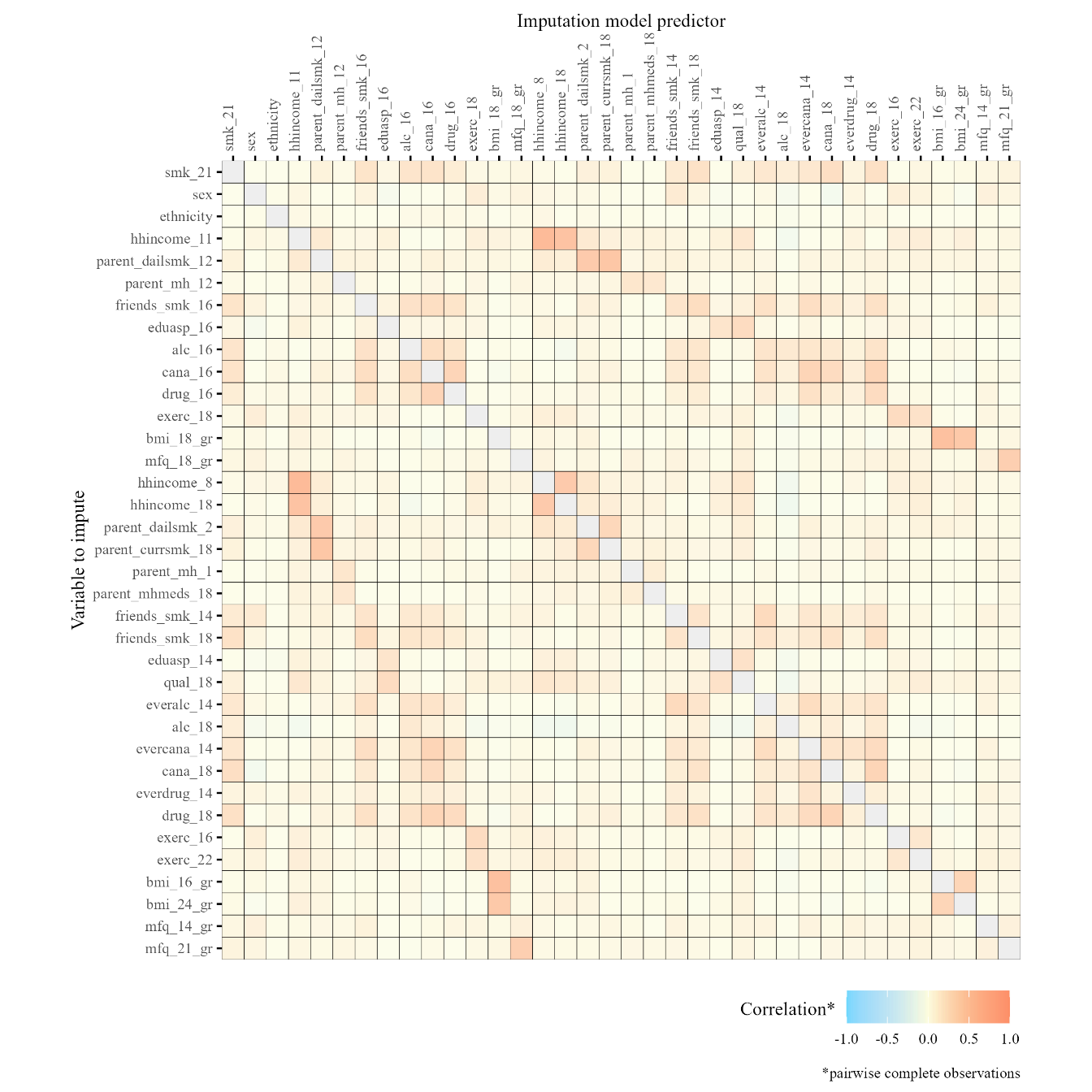

Supplementary Figure S3. Correlation matrix showing relationships between variables used in imputation within the eligible sample of participants who responded at the 21+ questionnaire (n=3,290) used to derive weights related to selection via smoking

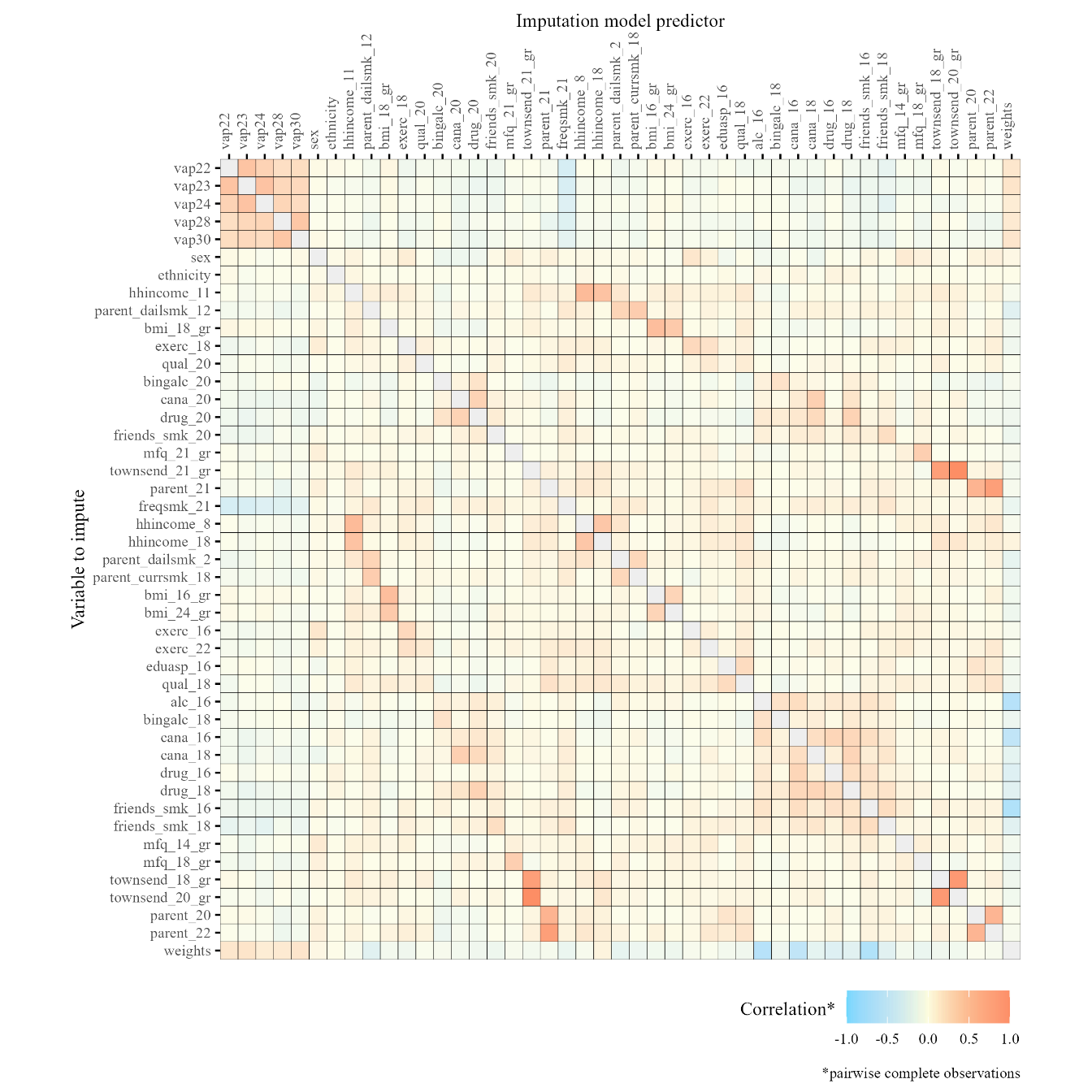

Supplementary Figure S4. Correlation matrix showing relationships between variables used in imputation within the analytic sample of people who smoked in the past 30 days at the 21+ questionnaire (n=858) used to investigate associations between participant characteristics and transitions from smoking using discrete time subdistribution hazard models

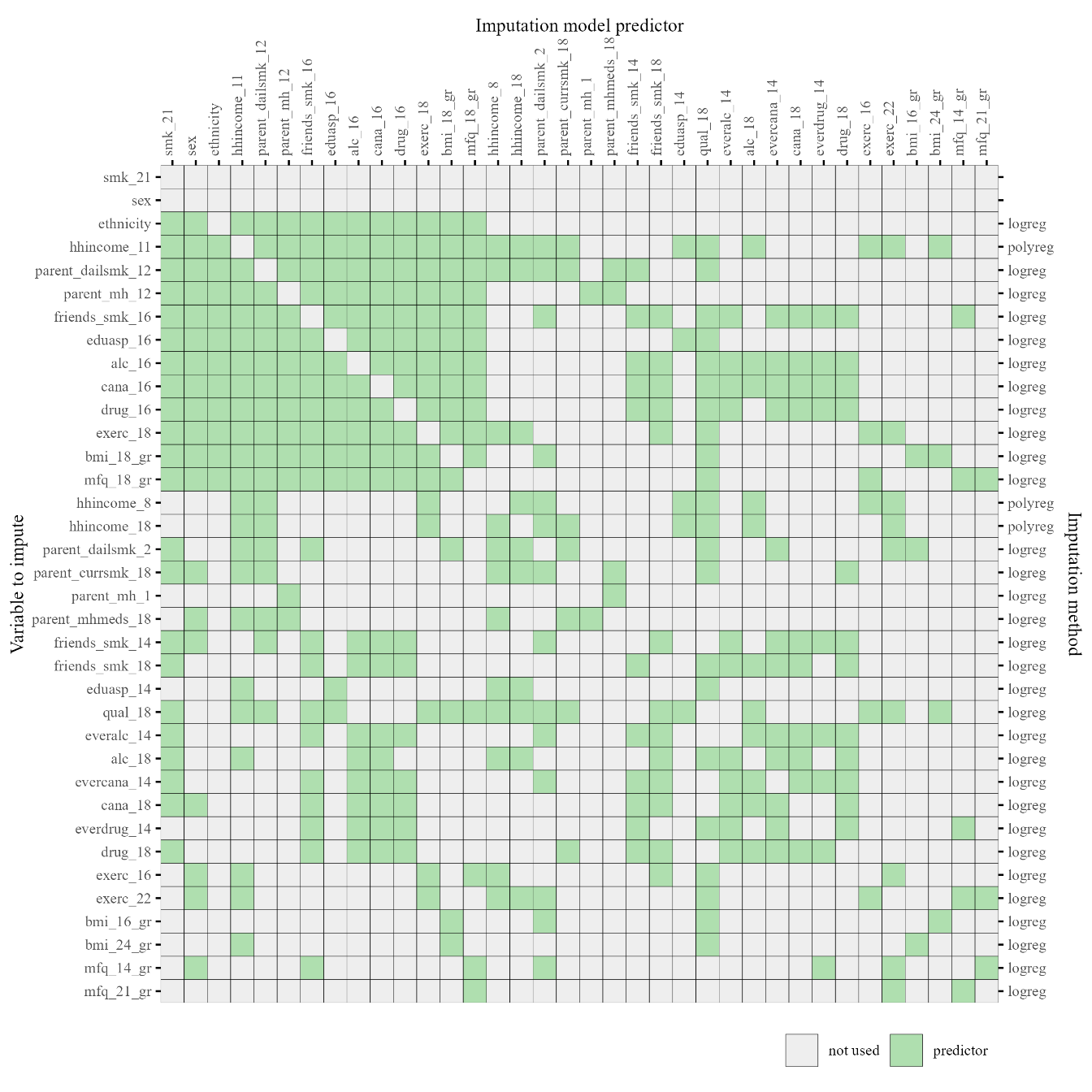

Supplementary Figure S5. Predictor matrix showing variables used in imputation within the eligible sample of participants who responded at the 21+ questionnaire (n=3,290) to derive weights related to selection via smoking

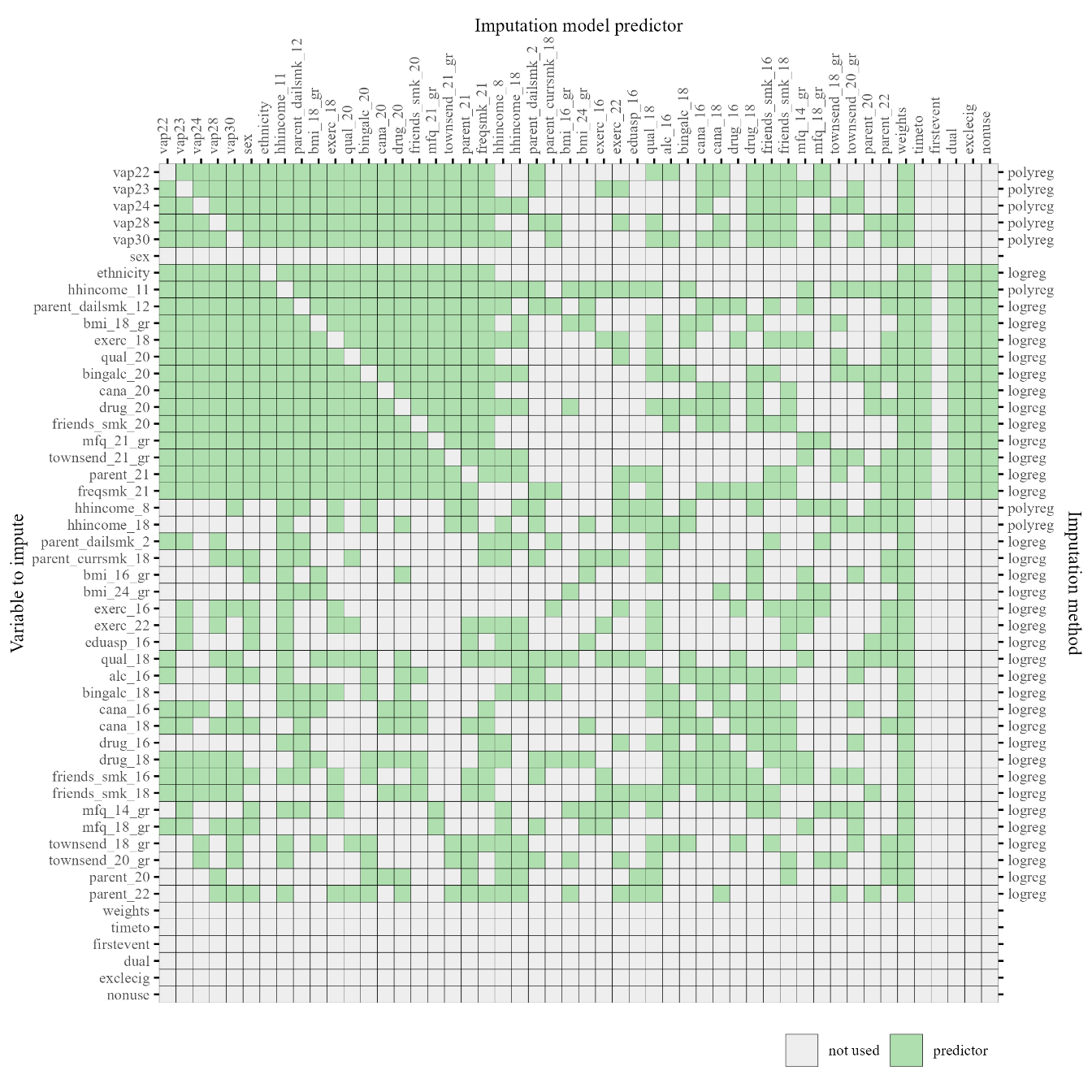

Supplementary Figure S6. Predictor matrix showing variables used in imputation within the analytic sample of people who smoked in the past 30 days at the 21+ questionnaire (n=858) used to investigate associations between participant characteristics and transitions from smoking using discrete time subdistribution hazard models

Supplementary Text S2. Description of approach used to address potential bias induced by conditioning sample on smoking

Collider bias occurs when an analysis controls for, stratifies on, or selects its sample based on a variable that is caused by both the exposure and the outcome. This distorts the association between the exposure and outcome and can lead to incorrect inferences from effect estimates^1^. In this case there are shared risk factors related to both e-cigarette and smoking (Supplementary Figure S1A). Therefore when selecting our sample to just those who smoke, we may induce a negative association between smoking risk factors and vaping and incorrectly conclude that risk factors for smoking are protective against vaping even though in reality they are unrelated^2^.

To mitigate this bias a two-step approach was utilised where first a logistic regression model is fitted to predict smoking in the past 30 days at age 21 (Supplementary Figure S5B). Secondly the regression model for e-cigarette use, estimated on the selected subsample of participants who smoke, is weighted using inverse predicted probabilities derived from the first model (Supplementary Figure S5C) to reduce bias^3^.

**References**

1.	Digitale JC, Martin JN, Glidden DV, Glymour MM. Key concepts in clinical epidemiology: collider-conditioning bias. *J Clin Epidemiol*. 2023;161:152-156. doi:10.1016/j.jclinepi.2023.07.004

2.	Tattan-Birch H, Marsden J, West R, Gage SH. Assessing and addressing collider bias in addiction research: the curious case of smoking and COVID-19. *Addiction*. 2021;116(5):982-984. doi:10.1111/add.15348

3.	Breen R, Ermisch J. Using Inverse Probability Weighting to Address Post-Outcome Collider Bias. *Sociol Methods Res*. 2024;53(1):5-27. doi:10.1177/00491241211043131

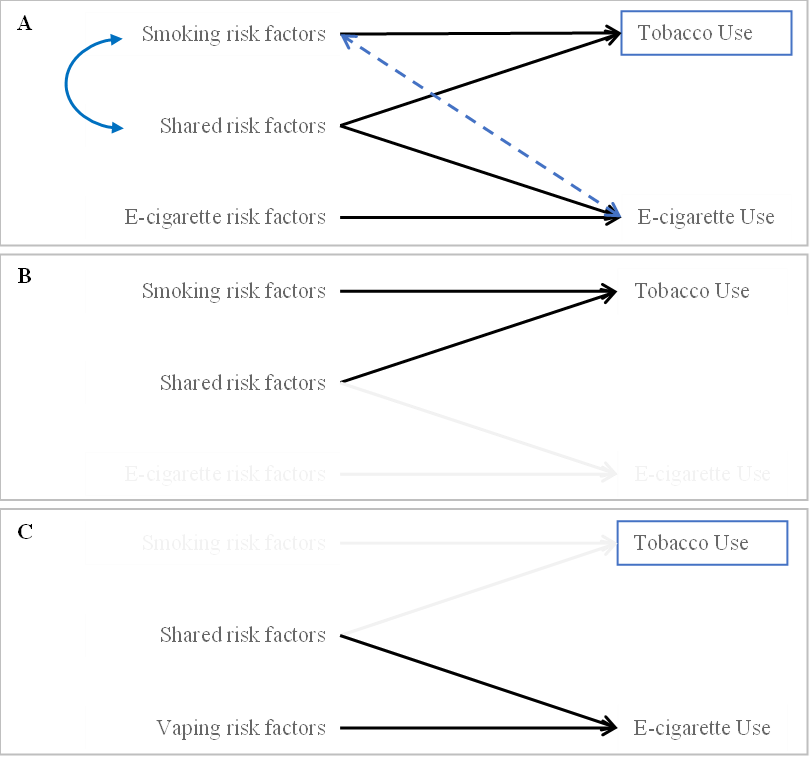

Supplementary Figure S7. Possible collider-conditioning bias when investigating e-cigarette use after conditioning on smoking and approach used to mitigate this bias

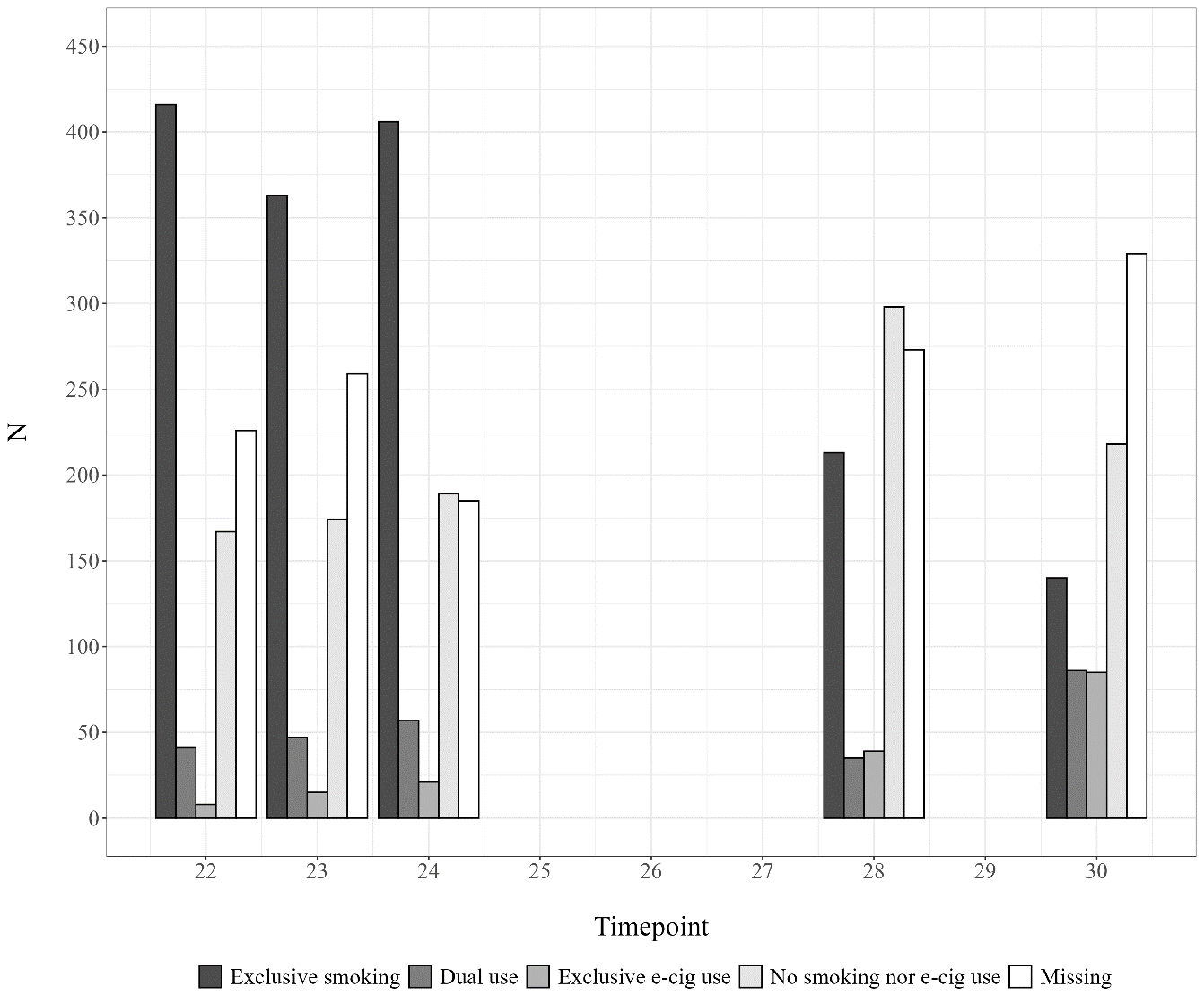

Supplementary Figure S8. Number of participants by self-reported nicotine use and missingness at each timepoint in the analytic sample (n=858)

Supplementary Table S1. Differences in characteristics by first reported transitions from smoking in those not loss to follow up, where transitions were derived by assuming transitions did not occur during any missing prior or intermediate reports of nicotine use, excluding any missingness in each characteristic

| **Characteristic** | **Stopping nicotine use** | | | **Exclusive e-cig use** | | | **Dual use** | | |
| --- | --- | --- | --- | --- | --- | --- | --- | --- | --- |
|  | **No*^1^*** | **Yes*^1^*** | **p***^2^* | **No*^1^*** | **Yes*^1^*** | **p***^3^* | **No*^1^*** | **Yes*^1^*** | **p***^2^* |
| ***Early-life confounders*** | | | | | | | | | |
| **Sex assigned at birth** |  |  | 0.093 |  |  | 0.009 |  |  | 0.019 |
| Male | 155 (37%) | 123 (32%) |  | 267 (36%) | 11 (19%) |  | 210 (33%) | 68 (43%) |  |
| Female | 260 (63%) | 265 (68%) |  | 478 (64%) | 47 (81%) |  | 433 (67%) | 92 (58%) |  |
| **Ethnicity** | *** | | | | | | | | |
| White |  |  |  |  |  |  |  |  |  |
| Minority ethnic |  |  |  |  |  |  |  |  |  |
| **Household income (11y)** |  |  | 0.700 |  |  | 0.800 |  |  | 0.400 |
| 560+ | 124 (39%) | 129 (44%) |  | 233 (41%) | 20 (43%) |  | 202 (41%) | 51 (41%) |  |
| 430-559 | 68 (22%) | 65 (22%) |  | 121 (21%) | 12 (26%) |  | 111 (23%) | 22 (18%) |  |
| 240-429 | 86 (27%) | 74 (25%) |  | 150 (27%) | 10 (21%) |  | 121 (25%) | 39 (32%) |  |
| <240 | 36 (11%) | 28 (9.5%) |  | 59 (10%) | 5 (11%) |  | 53 (11%) | 11 (8.9%) |  |
| **Parental smoking (12y)** |  |  | <0.001 |  |  | 0.500 |  |  | 0.007 |
| No | 239 (69%) | 272 (83%) |  | 475 (76%) | 36 (72%) |  | 418 (78%) | 93 (67%) |  |
| Yes | 107 (31%) | 57 (17%) |  | 150 (24%) | 14 (28%) |  | 118 (22%) | 46 (33%) |  |
| ***Substance use*** | | | | | | | | | |
| **Smoking frequency (21y)** |  |  | <0.001 |  |  | 0.003 |  |  | <0.001 |
| Occasional | 70 (17%) | 225 (58%) |  | 285 (38%) | 10 (18%) |  | 263 (41%) | 32 (20%) |  |
| Weekly or more | 341 (83%) | 162 (42%) |  | 458 (62%) | 45 (82%) |  | 375 (59%) | 128 (80%) |  |
| **Binge drinking frequency (20y)** |  |  | 0.600 |  |  | 0.800 |  |  | >0.9 |
| Never, monthly, or less | 173 (57%) | 164 (55%) |  | 314 (56%) | 23 (55%) |  | 271 (56%) | 66 (56%) |  |
| Weekly or more | 129 (43%) | 134 (45%) |  | 244 (44%) | 19 (45%) |  | 211 (44%) | 52 (44%) |  |
| **Cannabis use (20y)** |  |  | <0.001 |  |  | 0.800 |  |  | 0.700 |
| Never, less than monthly, or not in past year | 216 (70%) | 252 (83%) |  | 436 (76%) | 32 (78%) |  | 374 (76%) | 94 (78%) |  |
| Monthly or more | 92 (30%) | 53 (17%) |  | 136 (24%) | 9 (22%) |  | 118 (24%) | 27 (22%) |  |
| **Drug use (20y)** |  |  | 0.009 |  |  | 0.200 |  |  | 0.800 |
| No | 151 (51%) | 180 (62%) |  | 312 (57%) | 19 (46%) |  | 266 (56%) | 65 (58%) |  |
| Yes | 143 (49%) | 110 (38%) |  | 231 (43%) | 22 (54%) |  | 205 (44%) | 48 (42%) |  |
| ***Social and sociodemographic factors*** | | | | | | | | | |
| **Peer smoking (20y)** |  |  | <0.001 |  |  | 0.009 |  |  | 0.034 |
| None, a few, or some | 98 (32%) | 149 (49%) |  | 238 (42%) | 9 (21%) |  | 209 (43%) | 38 (32%) |  |
| Most or all | 205 (68%) | 158 (51%) |  | 330 (58%) | 33 (79%) |  | 282 (57%) | 81 (68%) |  |
| **Education (20y)** |  |  | <0.001 |  |  | 0.500 |  |  | 0.044 |
| Degree-level | 46 (15%) | 81 (26%) |  | 117 (20%) | 10 (24%) |  | 110 (22%) | 17 (14%) |  |
| A-level or lower/Other | 260 (85%) | 231 (74%) |  | 460 (80%) | 31 (76%) |  | 386 (78%) | 105 (86%) |  |
| **Being a parent (21y)** |  |  | 0.006 |  |  | 0.800 |  |  | 0.400 |
| No | 367 (90%) | 365 (95%) |  | 680 (92%) | 52 (91%) |  | 590 (92%) | 142 (90%) |  |
| Yes | 43 (10%) | 20 (5.2%) |  | 58 (7.9%) | 5 (8.8%) |  | 48 (7.5%) | 15 (9.6%) |  |
| **Neighbourhood deprivation (21y)** |  |  | 0.200 |  |  | 0.800 |  |  | 0.700 |
| Least deprived quintile | 133 (33%) | 143 (38%) |  | 258 (35%) | 18 (34%) |  | 219 (35%) | 57 (36%) |  |
| More deprived quintiles | 271 (67%) | 237 (62%) |  | 473 (65%) | 35 (66%) |  | 408 (65%) | 100 (64%) |  |
| ***Physical and mental health*** | | | | | | | | | |
| **BMI*^4^* (18y)** |  |  | 0.200 |  |  | 0.200 |  |  | 0.018 |
| <25 | 219 (75%) | 232 (80%) |  | 417 (77%) | 34 (85%) |  | 369 (80%) | 82 (69%) |  |
| >=25 | 72 (25%) | 58 (20%) |  | 124 (23%) | 6 (15%) |  | 94 (20%) | 36 (31%) |  |
| **Exercise frequency (18y)** |  |  | 0.010 |  |  | 0.120 |  |  | 0.800 |
| Weekly or more | 128 (59%) | 176 (70%) |  | 286 (66%) | 18 (53%) |  | 246 (65%) | 58 (64%) |  |
| Less than weekly | 89 (41%) | 74 (30%) |  | 147 (34%) | 16 (47%) |  | 130 (35%) | 33 (36%) |  |
| **Depressive symptoms (21y)** |  |  | 0.200 |  |  | 0.500 |  |  | 0.300 |
| <12 SFMQ*^5^* score | 312 (77%) | 302 (81%) |  | 570 (79%) | 44 (76%) |  | 494 (80%) | 120 (76%) |  |
| >=12 SFMQ score | 91 (23%) | 70 (19%) |  | 147 (21%) | 14 (24%) |  | 124 (20%) | 37 (24%) |  |
| *^1^* n (%);*^2^* Pearson's Chi-squared test; *^3^* Pearson's Chi-squared test; Fisher's exact test; *^4^* BMI = Body Mass Index; *^5^* SMFQ = Short Mood and Feelings Questionnaire; *** Omitted to avoid identification of small cell counts (<5) due to the low number of participants from a minority ethnic background | | | | | | | | | |

Supplementary Table S2. Differences in characteristics by early-life confounders excluding any pairwise missingness

|  | **Sex assigned at birth** | | | **Household income** | | | | | **Parental smoking** | | |
| --- | --- | --- | --- | --- | --- | --- | --- | --- | --- | --- | --- |
| **Characteristic** | **Male***^1^* | **Female***^1^* | **p***^2^* | **560+***^1^* | **430-559***^1^* | **240-429***^1^* | **<240***^1^* | **p***^3^* | **No***^1^* | **Yes***^1^* | **p***^2^* |
| ***Early-life confounders*** | | | | | | | | | | | |
| **Sex assigned at birth** |  |  |  |  |  |  |  | 0.140 |  |  | 0.900 |
| Male |  |  |  | 118 (45%) | 53 (38%) | 64 (36%) | 22 (32%) |  | 210 (38%) | 68 (39%) |  |
| Female |  |  |  | 147 (55%) | 88 (62%) | 112 (64%) | 47 (68%) |  | 337 (62%) | 106 (61%) |  |
| **Ethnicity** | *** | | | | | | | | | | |
| White |  |  |  |  |  |  |  |  |  |  |  |
| Minority ethnic |  |  |  |  |  |  |  |  |  |  |  |
| **Household income (11y)** |  |  | 0.140 |  |  |  |  |  |  |  | 0.009 |
| 560+ | 118 (46%) | 147 (37%) |  |  |  |  |  |  | 211 (45%) | 42 (31%) |  |
| 430-559 | 53 (21%) | 88 (22%) |  |  |  |  |  |  | 100 (21%) | 31 (23%) |  |
| 240-429 | 64 (25%) | 112 (28%) |  |  |  |  |  |  | 120 (26%) | 44 (32%) |  |
| <240 | 22 (8.6%) | 47 (12%) |  |  |  |  |  |  | 38 (8.1%) | 20 (15%) |  |
| **Parental smoking (12y)** |  |  | 0.900 |  |  |  |  | 0.009 |  |  |  |
| No | 210 (76%) | 337 (76%) |  | 211 (83%) | 100 (76%) | 120 (73%) | 38 (66%) |  |  |  |  |
| Yes | 68 (24%) | 106 (24%) |  | 42 (17%) | 31 (24%) | 44 (27%) | 20 (34%) |  |  |  |  |
| ***Substance use*** | | | | | | | | | | | |
| **Smoking frequency (21y)** |  |  | 0.300 |  |  |  |  | 0.079 |  |  | <0.001 |
| Occasional | 120 (39%) | 192 (35%) |  | 111 (42%) | 51 (36%) | 57 (33%) | 19 (28%) |  | 236 (43%) | 36 (21%) |  |
| Weekly or more | 187 (61%) | 354 (65%) |  | 153 (58%) | 89 (64%) | 116 (67%) | 50 (72%) |  | 307 (57%) | 137 (79%) |  |
| **Binge drinking frequency (20y)** |  |  | <0.001 |  |  |  |  | 0.016 |  |  | 0.081 |
| Never, monthly, or less | 100 (44%) | 253 (63%) |  | 99 (47%) | 53 (52%) | 72 (63%) | 33 (65%) |  | 216 (53%) | 78 (62%) |  |
| Weekly or more | 126 (56%) | 151 (37%) |  | 111 (53%) | 48 (48%) | 42 (37%) | 18 (35%) |  | 191 (47%) | 48 (38%) |  |
| **Cannabis use (20y)** |  |  | <0.001 |  |  |  |  | 0.093 |  |  | 0.150 |
| Never, less than monthly, or not past year | 150 (66%) | 339 (82%) |  | 148 (71%) | 84 (81%) | 96 (79%) | 34 (67%) |  | 321 (78%) | 91 (72%) |  |
| Monthly or more | 78 (34%) | 76 (18%) |  | 61 (29%) | 20 (19%) | 26 (21%) | 17 (33%) |  | 91 (22%) | 36 (28%) |  |
| **Drug use (20y)** |  |  | <0.001 |  |  |  |  | 0.700 |  |  | 0.700 |
| No | 100 (46%) | 247 (62%) |  | 108 (53%) | 55 (55%) | 67 (60%) | 28 (57%) |  | 223 (57%) | 66 (55%) |  |
| Yes | 116 (54%) | 150 (38%) |  | 97 (47%) | 45 (45%) | 45 (40%) | 21 (43%) |  | 170 (43%) | 55 (45%) |  |
| ***Social and sociodemographic factors*** | | | | | | | | | | | |
| **Peer smoking (20y)** |  |  | 0.200 |  |  |  |  | >0.9 |  |  | 0.100 |
| None, a few, or some | 99 (44%) | 162 (39%) |  | 87 (41%) | 42 (41%) | 48 (39%) | 19 (37%) |  | 180 (44%) | 45 (35%) |  |
| Most or all | 126 (56%) | 253 (61%) |  | 123 (59%) | 60 (59%) | 74 (61%) | 32 (63%) |  | 232 (56%) | 82 (65%) |  |
| **Education (20y)** |  |  | 0.900 |  |  |  |  | 0.5 |  |  | 0.200 |
| Degree-level | 47 (20%) | 87 (21%) |  | 50 (23%) | 28 (27%) | 26 (22%) | 8 (15%) |  | 99 (24%) | 23 (18%) |  |
| A-level or lower/Other | 184 (80%) | 329 (79%) |  | 165 (77%) | 77 (73%) | 93 (78%) | 44 (85%) |  | 317 (76%) | 103 (82%) |  |
| **Being a parent (21y)** |  |  | <0.001 |  |  |  |  | <0.001 |  |  | 0.6 |
| No | 293 (96%) | 486 (89%) |  | 255 (97%) | 132 (95%) | 154 (88%) | 62 (91%) |  | 499 (92%) | 163 (94%) |  |
| Yes | 11 (3.6%) | 59 (11%) |  | 7 (2.7%) | 7 (5.0%) | 21 (12%) | 6 (8.8%) |  | 41 (7.6%) | 11 (6.3%) |  |
| **Neighbourhood deprivation (21y)** |  |  | 0.120 |  |  |  |  | <0.001 |  |  | 0.020 |
| Least deprived quintile | 114 (39%) | 179 (33%) |  | 114 (44%) | 49 (36%) | 48 (28%) | 14 (21%) |  | 205 (38%) | 48 (29%) |  |
| More deprived quintiles | 182 (61%) | 360 (67%) |  | 144 (56%) | 87 (64%) | 124 (72%) | 54 (79%) |  | 328 (62%) | 120 (71%) |  |
| ***Physical and mental health*** | | | | | | | | | | | |
| **BMI*^4^* (18y)** |  |  | 0.041 |  |  |  |  | 0.006 |  |  | 0.400 |
| <25 | 192 (82%) | 278 (75%) |  | 174 (86%) | 85 (77%) | 91 (71%) | 27 (69%) |  | 331 (80%) | 91 (76%) |  |
| >=25 | 42 (18%) | 93 (25%) |  | 29 (14%) | 26 (23%) | 37 (29%) | 12 (31%) |  | 84 (20%) | 29 (24%) |  |
| **Exercise frequency (18y)** |  |  | <0.001 |  |  |  |  | 0.047 |  |  | 0.083 |
| Weekly or more | 124 (76%) | 190 (60%) |  | 120 (74%) | 44 (62%) | 61 (59%) | 25 (63%) |  | 227 (68%) | 52 (58%) |  |
| Less than weekly | 40 (24%) | 129 (40%) |  | 42 (26%) | 27 (38%) | 43 (41%) | 15 (38%) |  | 109 (32%) | 38 (42%) |  |
| **Depressive symptoms (21y)** |  |  | <0.001 |  |  |  |  | >0.9 |  |  | 0.500 |
| <12 SFMQ*^5^* score | 256 (86%) | 397 (75%) |  | 205 (80%) | 106 (79%) | 136 (79%) | 53 (79%) |  | 420 (80%) | 139 (82%) |  |
| >=12 SFMQ score | 40 (14%) | 135 (25%) |  | 52 (20%) | 29 (21%) | 37 (21%) | 14 (21%) |  | 107 (20%) | 30 (18%) |  |
| *^1^* n (%); *^2^* Pearson's Chi-squared test; *^3^* Pearson's Chi-squared test; Fisher's exact test; *^4^* BMI = Body Mass Index; *^5^* SMFQ = Short Mood and Feelings Questionnaire; *** Omitted to avoid identification of small cell counts (<5) due to the low number of participants from a minority ethnic background | | | | | | | | | | | |

Supplementary Table S3. Pooled summary statistics for *early-life confounders* and time from sub-distribution discrete time survival analyses, weighted for selection via smoking

|  | **Stopping nicotine use (52%)** | | | **Exclusive e-cig use (9%)** | | | **Dual use (27%)** | | |
| --- | --- | --- | --- | --- | --- | --- | --- | --- | --- |
| **Characteristic** | **SHR***^3^* | **95% CI***^1^* | **p** | **SHR***^3^* | **95% CI***^1^* | **p** | **SHR***^3^* | **95% CI***^1^* | **p** |
| **Years since 21+ questionnaire*^4^*** | 0.72 | 0.65, 0.79 | <0.001 | 1.05 | 0.82, 1.36 | 0.7 | 0.87 | 0.76, 0.99 | 0.04 |
| **Sex assigned at birth** |  |  |  |  |  |  |  |  |  |
| *Male* | — | — |  | — | — |  | — | — |  |
| *Female* | 1.25 | 0.98, 1.59 | 0.07 | 1.62 | 0.75, 3.49 | 0.2 | 0.69 | 0.49, 0.98 | 0.036 |
| **Ethnicity** |  |  |  |  |  |  |  |  |  |
| *White* | — | — |  | — | — |  | — | — |  |
| *Minority ethnic* | 1.3 | 0.72, 2.37 | 0.4 | 0.01 | 0.00, 4,582 | 0.5 | 0.71 | 0.25, 2.05 | 0.5 |
| **Household income (11y)** |  |  |  |  |  |  |  |  |  |
| *560+* | — | — |  | — | — |  | — | — |  |
| *430-559* | 0.98 | 0.71, 1.35 | >0.9 | 0.97 | 0.42, 2.23 | >0.9 | 0.8 | 0.49, 1.31 | 0.4 |
| *240-429* | 0.95 | 0.71, 1.27 | 0.7 | 0.96 | 0.42, 2.17 | >0.9 | 1.19 | 0.78, 1.80 | 0.4 |
| *<240* | 0.8 | 0.53, 1.20 | 0.3 | 0.89 | 0.28, 2.83 | 0.8 | 0.93 | 0.49, 1.77 | 0.8 |
| **Parental smoking (12y)** |  |  |  |  |  |  |  |  |  |
| *No* | — | — |  | — | — |  | — | — |  |
| *Yes* | 0.6 | 0.44, 0.81 | 0.001 | 1.27 | 0.63, 2.57 | 0.5 | 1.58 | 1.08, 2.32 | 0.02 |
| *^1^* SHR = Sub-distribution Hazard Ratio; *^2^* Discrete-time indicator where coefficients represent the effect of a one-unit increase in the time interval on the sub-distribution hazard ratio | | | | | | | | | |

Supplementary Table S4. Pooled summary statistics from sub-distribution discrete time survival analyses, adjusted for early-life confounders, *unweighted*

| **Characteristic** | **Stopping nicotine use (52%)** | | | **Exclusive e-cig use (9%)** | | | **Dual use (27%)** | | |
| --- | --- | --- | --- | --- | --- | --- | --- | --- | --- |
|  | **SHR***^1^* | **95% CI***^1^* | **p** | **SHR***^1^* | **95% CI***^1^* | **p** | **SHR***^1^* | **95% CI***^1^* | **p** |
| ***Substance use*** | | | | | | | | | |
| **Smoking frequency (21y)** |  |  |  |  |  |  |  |  |  |
| Occasional | — | — |  | — | — |  | — | — |  |
| Weekly or more | 0.28 | 0.22, 0.34 | <0.001 | 2.64 | 1.27, 5.47 | 0.009 | 2.60 | 1.75, 3.87 | <0.001 |
| **Binge drinking frequency (20y)** |  |  |  |  |  |  |  |  |  |
| Never, monthly, or less | — | — |  | — | — |  | — | — |  |
| Weekly or more | 1.03 | 0.81, 1.30 | 0.8 | 1.16 | 0.62, 2.18 | 0.6 | 0.92 | 0.65, 1.30 | 0.6 |
| **Cannabis use (20y)** |  |  |  |  |  |  |  |  |  |
| Never, less than monthly, not past year | — | — |  | — | — |  | — | — |  |
| Monthly or more | 0.65 | 0.48, 0.87 | 0.004 | 1.01 | 0.48, 2.14 | >0.9 | 1.15 | 0.79, 1.66 | 0.5 |
| **Drug use (20y)** |  |  |  |  |  |  |  |  |  |
| No | — | — |  | — | — |  | — | — |  |
| Yes | 0.75 | 0.59, 0.95 | 0.015 | 1.36 | 0.75, 2.46 | 0.3 | 1.03 | 0.74, 1.45 | 0.8 |
| ***Social and sociodemographic factors*** | | | | | | | | | |
| **Peer smoking (20y)** |  |  |  |  |  |  |  |  |  |
| None, a few, or some | — | — |  | — | — |  | — | — |  |
| Most or all | 0.62 | 0.50, 0.78 | <0.001 | 1.82 | 0.89, 3.74 | 0.10 | 1.43 | 1.01, 2.04 | 0.046 |
| **Education (20y)** |  |  |  |  |  |  |  |  |  |
| Degree-level | — | — |  | — | — |  | — | — |  |
| A-level or lower/Other | 0.68 | 0.52, 0.88 | 0.004 | 0.72 | 0.36, 1.42 | 0.3 | 1.64 | 1.02, 2.66 | 0.043 |
| **Being a parent (21y)** |  |  |  |  |  |  |  |  |  |
| No | — | — |  | — | — |  | — | — |  |
| Yes | 0.51 | 0.32, 0.83 | 0.006 | 1.08 | 0.37, 3.13 | 0.9 | 1.28 | 0.76, 2.15 | 0.3 |
| **Neighbourhood deprivation (21y)** |  |  |  |  |  |  |  |  |  |
| Least deprived quintile | — | — |  | — | — |  | — | — |  |
| More deprived quintiles | 0.91 | 0.73, 1.14 | 0.4 | 0.95 | 0.52, 1.75 | 0.9 | 0.97 | 0.70, 1.33 | 0.8 |
| ***Physical and mental health*** | | | | | | | | | |
| **BMI*^2^* (18y)** |  |  |  |  |  |  |  |  |  |
| <25 | — | — |  | — | — |  | — | — |  |
| >=25 | 0.91 | 0.68, 1.20 | 0.5 | 0.63 | 0.26, 1.48 | 0.3 | 1.59 | 1.11, 2.27 | 0.011 |
| **Exercise frequency (18y)** |  |  |  |  |  |  |  |  |  |
| Weekly or more | — | — |  | — | — |  | — | — |  |
| Less than weekly | 0.71 | 0.54, 0.94 | 0.018 | 1.20 | 0.60, 2.40 | 0.6 | 1.10 | 0.75, 1.61 | 0.6 |
| **Depressive symptoms (21y)** |  |  |  |  |  |  |  |  |  |
| <12 SMFQ*^3^* score | — | — |  | — | — |  | — | — |  |
| >=12 SMFQ score | 0.79 | 0.61, 1.03 | 0.079 | 1.12 | 0.59, 2.11 | 0.7 | 1.14 | 0.81, 1.63 | 0.4 |
| *^1^* SHR = Sub-distribution Hazard Ratio, CI = Confidence Interval; *^2^* BMI = Body Mass Index; *^3^* SMFQ = Short Mood and Feelings Questionnaire | | | | | | | | | |

Supplementary Table S5. Pooled summary statistics from sub-distribution discrete time survival analyses, *unadjusted*, weighted for selection via smoking

| **Characteristic** | **Stopping nicotine use (52%)** | | | **Exclusive e-cig use (9%)** | | | **Dual use (27%)** | | |
| --- | --- | --- | --- | --- | --- | --- | --- | --- | --- |
|  | **SHR***^1^* | **95% CI***^1^* | **p** | **SHR***^1^* | **95% CI***^1^* | **p** | **SHR***^1^* | **95% CI***^1^* | **p** |
| ***Substance use*** | | | | | | | | | |
| **Smoking frequency (21y)** |  |  |  |  |  |  |  |  |  |
| Occasional | — | — |  | — | — |  | — | — |  |
| Weekly or more | 0.28 | 0.22, 0.35 | <0.001 | 2.92 | 1.31, 6.49 | 0.009 | 3.03 | 1.98, 4.63 | <0.001 |
| **Binge drinking (20y)** |  |  |  |  |  |  |  |  |  |
| Never, monthly, or less | — | — |  | — | — |  | — | — |  |
| Weekly or more | 1.01 | 0.79, 1.29 | >0.9 | 1.10 | 0.56, 2.16 | 0.8 | 0.95 | 0.66, 1.37 | 0.8 |
| **Cannabis use (20y)** |  |  |  |  |  |  |  |  |  |
| Never, less than monthly, not past year | — | — |  | — | — |  | — | — |  |
| Monthly or more | 0.64 | 0.47, 0.87 | 0.005 | 0.98 | 0.45, 2.14 | >0.9 | 1.27 | 0.85, 1.90 | 0.3 |
| **Drug use (20y)** |  |  |  |  |  |  |  |  |  |
| No | — | — |  | — | — |  | — | — |  |
| Yes | 0.76 | 0.60, 0.98 | 0.032 | 1.35 | 0.71, 2.58 | 0.4 | 1.11 | 0.78, 1.60 | 0.6 |
| ***Social and sociodemographic factors*** | | | | | | | | | |
| **Peer smoking (20y)** |  |  |  |  |  |  |  |  |  |
| None, a few, or some | — | — |  | — | — |  | — | — |  |
| Most or all | 0.64 | 0.51, 0.81 | <0.001 | 1.76 | 0.83, 3.70 | 0.14 | 1.53 | 1.06, 2.19 | 0.022 |
| **Education (20y)** |  |  |  |  |  |  |  |  |  |
| Degree-level | — | — |  | — | — |  | — | — |  |
| A-level or lower/Other | 0.68 | 0.51, 0.89 | 0.005 | 0.66 | 0.32, 1.37 | 0.3 | 1.77 | 1.05, 2.96 | 0.031 |
| **Being a parent (21y)** |  |  |  |  |  |  |  |  |  |
| No | — | — |  | — | — |  | — | — |  |
| Yes | 0.49 | 0.29, 0.82 | 0.007 | 1.37 | 0.44, 4.27 | 0.6 | 1.46 | 0.86, 2.50 | 0.2 |
| **Neighbourhood deprivation (21y)** |  |  |  |  |  |  |  |  |  |
| Least deprived quintile | — | — |  | — | — |  | — | — |  |
| More deprived quintiles | 0.90 | 0.72, 1.13 | 0.4 | 1.06 | 0.54, 2.07 | 0.9 | 1.01 | 0.72, 1.43 | >0.9 |
| ***Physical and mental health*** | | | | | | | | | |
| **BMI*^2^* (18y)** |  |  |  |  |  |  |  |  |  |
| <25 | — | — |  | — | — |  | — | — |  |
| >=25 | 0.88 | 0.66, 1.16 | 0.3 | 0.67 | 0.27, 1.64 | 0.4 | 1.56 | 1.07, 2.26 | 0.019 |
| **Frequency of exercise, past year (18y)** |  |  |  |  |  |  |  |  |  |
| Weekly or more | — | — |  | — | — |  | — | — |  |
| Less than weekly | 0.73 | 0.55, 0.97 | 0.030 | 1.40 | 0.68, 2.89 | 0.4 | 1.03 | 0.68, 1.55 | 0.9 |
| **Depressive symptoms (21y)** |  |  |  |  |  |  |  |  |  |
| <12 SMFQ*^3^* score | — | — |  | — | — |  | — | — |  |
| >=12 SMFQ score | 0.89 | 0.68, 1.17 | 0.4 | 1.20 | 0.61, 2.37 | 0.6 | 1.15 | 0.79, 1.68 | 0.5 |
| *^1^* SHR = Sub-distribution Hazard Ratio, CI = Confidence Interval; *^2^* BMI = Body Mass Index; *^3^* SMFQ = Short Mood and Feelings Questionnaire | | | | | | | | | |

Supplementary Table S6. Summary statistics from sub-distribution discrete time survival, adjusted for early-life confounders, *complete cases* (n=168), unweighted (as weights rely on imputed data)

| **Characteristic** | **Stopping nicotine use (n=90)** | | | **Exclusive e-cig use (n=12)** | | | **Dual use (n=36)** | | |
| --- | --- | --- | --- | --- | --- | --- | --- | --- | --- |
|  | **SHR***^1^* | **95% CI***^1^* | **p** | **SHR***^1^* | **95% CI***^1^* | **p** | **SHR***^1^* | **95% CI***^1^* | **p** |
| ***Substance use*** | | | | | | | | | |
| **Smoking frequency (21y)** |  |  |  |  |  |  |  |  |  |
| Occasional | — | — |  | — | — |  | — | — |  |
| Weekly or more | 0.25 | 0.16, 0.40 | <0.001 | 3.24 | 0.93, 14.9 | 0.085 | 2.27 | 1.12, 4.90 | 0.028 |
| **Binge drinking (20y)** |  |  |  |  |  |  |  |  |  |
| Never, monthly, or less | — | — |  | — | — |  | — | — |  |
| Weekly or more | 0.96 | 0.62, 1.49 | 0.9 | 2.10 | 0.63, 8.17 | 0.2 | 0.83 | 0.41, 1.65 | 0.6 |
| **Cannabis use (20y)** |  |  |  |  |  |  |  |  |  |
| Never, less than monthly, not past year | — | — |  | — | — |  | — | — |  |
| Monthly or more | 0.46 | 0.24, 0.83 | 0.014 | 1.45 | 0.37, 4.93 | 0.6 | 1.24 | 0.55, 2.57 | 0.6 |
| **Drug use (20y)** |  |  |  |  |  |  |  |  |  |
| No | — | — |  | — | — |  | — | — |  |
| Yes | 0.59 | 0.37, 0.93 | 0.023 | 1.77 | 0.55, 6.22 | 0.3 | 1.19 | 0.60, 2.34 | 0.6 |
| ***Social and sociodemographic factors*** | | | | | | | | | |
| **Peer smoking (20y)** |  |  |  |  |  |  |  |  |  |
| None, a few, or some | — | — |  | — | — |  | — | — |  |
| Most or all | 0.60 | 0.39, 0.94 | 0.022 | 1.18 | 0.36, 4.54 | 0.8 | 1.57 | 0.77, 3.38 | 0.2 |
| **Education (20y)** |  |  |  |  |  |  |  |  |  |
| Degree-level | — | — |  | — | — |  | — | — |  |
| A-level or lower/Other | 0.42 | 0.27, 0.68 | <0.001 | 1.63 | 0.41, 10.9 | 0.5 | 3.61 | 1.28, 15.1 | 0.035 |
| **Being a parent (21y)** |  |  |  |  |  |  |  |  |  |
| No | — | — |  | — | — |  | — | — |  |
| Yes | 0.00 |  | >0.9 | 0.00 |  | >0.9 | 4.98 | 1.05, 18.3 | 0.023 |
| **Neighbourhood deprivation (21y)** |  |  |  |  |  |  |  |  |  |
| Least deprived quintile | — | — |  | — | — |  | — | — |  |
| More deprived quintiles | 1.01 | 0.65, 1.57 | >0.9 | 0.92 | 0.27, 3.18 | 0.9 | 0.77 | 0.39, 1.54 | 0.5 |
| ***Physical and mental health*** | | | | | | | | | |
| **BMI^2^ (18y)** |  |  |  |  |  |  |  |  |  |
| <25 | — | — |  | — | — |  | — | — |  |
| >=25 | 0.87 | 0.46, 1.54 | 0.6 | 0.40 | 0.02, 2.15 | 0.4 | 1.45 | 0.60, 3.13 | 0.4 |
| **Exercise frequency (18y)** |  |  |  |  |  |  |  |  |  |
| Weekly or more | — | — |  | — | — |  | — | — |  |
| Less than weekly | 0.59 | 0.36, 0.93 | 0.028 | 1.62 | 0.47, 5.30 | 0.4 | 1.36 | 0.67, 2.68 | 0.4 |
| **Depressive symptoms (21y)** |  |  |  |  |  |  |  |  |  |
| <12 SMFQ^3^ score | — | — |  | — | — |  | — | — |  |
| >=12 SMFQ score | 0.78 | 0.41, 1.37 | 0.4 | 2.12 | 0.46, 7.48 | 0.3 | 1.11 | 0.41, 2.54 | 0.8 |
| *^1^* SHR = Sub-distribution Hazard Ratio, CI = Confidence Interval; *^2^* BMI = Body Mass Index; *^3^* SMFQ = Short Mood and Feelings Questionnaire | | | | | | | | | |
